# Using a population-based Kalman estimator to model the COVID-19 epidemic in France: estimating associations between disease transmission and non-pharmaceutical interventions

**DOI:** 10.1101/2021.07.09.21260259

**Authors:** Annabelle Collin, Boris P. Hejblum, Carole Vignals, Laurent Lehot, Rodolphe Thiébaut, Philippe Moireau, Mélanie Prague

**Affiliations:** Inria, Inria Bordeaux - Sud-Ouest, Bordeaux INP, IMB UMR 5251, IMB UMR 5251, Université Bordeaux, Talence, France; Inria, Inria Bordeaux - Sud-Ouest, Talence, Univ. Bordeaux, Inserm, Bordeaux Population Health Research Center, SISTM Team, UMR 1219, F-33000 Bordeaux, Vaccine Research Institute, F-94000 Créteil, France; Inria, Inria Bordeaux - Sud-Ouest, Talence, Univ. Bordeaux, Inserm, Bordeaux Population Health Research Center, SISTM Team, UMR 1219, F-33000 Bordeaux, Vaccine Research Institute, F-94000 Créteil, CHU Pellegrin, F-33000 Bordeaux, France; Inria, Inria Saclay-Ile-de-France, LMS, CNRS UMR 77 07, Ecole Polytechnique, Institut Polytechnique de Paris, France

**Keywords:** COVID-19, Epidemic modeling, Non-pharmaceutical interventions, Kalman filters, Population estimation

## Abstract

**Summary:** In response to the COVID-19 pandemic caused by SARS-CoV-2, governments have adopted a wide range of non-pharmaceutical interventions (NPI). These include stringent measures such as strict lockdowns, closing schools, bars and restaurants, curfews, and barrier gestures such as mask-wearing and social distancing. Deciphering the effectiveness of each NPI is critical to responding to future waves and outbreaks. To this end, we first develop a dynamic model of the French COVID-19 epidemics over a one-year period. We rely on a global extended Susceptible-Infectious-Recovered (SIR) mechanistic model of infection that includes a dynamic transmission rate over time. Multilevel data across French regions are integrated using random effects on the parameters of the mechanistic model, boosting statistical power by multiplying integrated observation series. We estimate the parameters using a new population-based statistical approach based on a Kalman filter, used for the first time in analysing real-world data. We then fit the estimated time-varying transmission rate using a regression model that depends on the NPIs while accounting for vaccination coverage, the occurrence of variants of concern (VoC), and seasonal weather conditions. We show that all NPIs considered have an independent significant association with transmission rates. In addition, we show a strong association between weather conditions that reduces transmission in summer, and we also estimate increased transmissibility of VoC.

## 1 Introduction

The World Health Organization declared a pandemic COVID-19 on March 11, 2020. This disease is caused by infection with the SARS-CoV-2 virus. By April 30, 2021, more than 150 million cases have been confirmed worldwide, including 3.16 million deaths. While the majority of infected cases have a mild form (upper respiratory tract infection symptoms) with no special care needs (1), about 3% of cases, especially the elderly, require hospitalization for treatment, such as oxygen therapy (2; 3; 4). About 17% of these cases are severe forms (severe acute respiratory syndrome) that require treatment in the intensive care units (ICU) and possibly mechanical ventilation (5).

The COVID-19 pandemic has stretched modern healthcare systems around the world to their limits. Because SARS-CoV-2 is an emerging pathogen, the entire human population is vulnerable to infection. Wherever there are outbreaks of SARS-CoV-2, there has been an increase in hospital admissions and especially in the need for intensive care units. Because of an infectious phase that begins before any symptoms are apparent and a significant proportion of a- or pauci-symptomatic infections (6), the spread of SARS-CoV-2 is particularly difficult to control (7). In response, most governments have taken drastic public health measures, also known as non-pharmaceutical interventions (NPI), to reduce the transmission of SARS-CoV-2 in their populations and thus reduce the pressure on their health systems. In particular, the French government has adopted the concept of a “graduated response” to the pandemic and has deployed an arsenal of different NPIs – some very stringent, others less stringent – in response to the COVID-19 national epidemic situation. Hale et al. (8) have created a stringency index that helps to understand how strong the measures have been over time. However, this indicator does not allow us to distinguish the effectiveness of each NPI, which is critical for future preparedness response plans. Because all NPIs have economic, psychological, and social costs, it is critical to assess their impact on SARS-CoV-2 transmission and on the dynamics of the COVID-19 epidemic.

Many studies relied on mechanistic models of epidemics to either predict progression (9), evaluate vaccine prioritization strategies (10), or retrospectively measure the impact of NPIs. Early in the epidemic, the focus was on the timing of NPI initiation (11; 12) rather than their impact. Disentangling the impact of individual NPIs is a complex problem, in particular because their allocation is not random and depends on the state of the epidemic. Many attempts aggregated data from multiple countries. Some worked with time-series regression based on incidence data (13; 14; 15; 16), with semi-mechanistic models and assessed the percentage reduction in the effective reproductive number (17; 18; 19; 20; 21; 75) or with advanced machine learning approaches (22). Instead, we limit ourselves to France, to avoid potential confusion bias in the effect estimation due to differences in behavior and adherence levels across various populations. Most of the work published at the country level focused on a single aggregate NPI such as the Oxford COVID-19 Government Response Tracker (8), a very early epidemic (23; 24), or a very limited number of interventions, see Brauner et al. for a review (25). Only a few works were interested in multiple waves of epidemics (26; 27; 28). In this study, as in only a few other works as for example Ge et al. (29), we aim to consider a rather long observation period (the first 12 months of the pandemic).

In France, although some studies have quantified the effect of several NPIs (3; 30), the authors base their results on the earliest phase of the epidemic and examine only a limited number of NPIs. Our work focuses instead on the impact of individual NPIs on the transmission rate. This is a better and more valid indicator than direct epidemic curves or the reproductive number, because it is independent of the proportion of the population infected. Finally, previous work does not take into account the prevalence of vaccination, the spread of new variants, and the importance of weather in estimating the impact of NPIs. Quantifying the associations between NPIs and transmission rate leads to a challenging estimation problem, including concerns about the practical identifiability of each effect.

In this work, we propose a two-step approach. First, we estimate the transmission rate of SARS-CoV-2 and its variations in the 12 non-island French regions over a period of more than one year - from March 2, 2020 to March 28, 2021. Second, we use linear regression to estimate the associations between different NPIs and transmission rates. We account for seasonal weather conditions during the pandemic, as well as the occurrence of non-historical variants of concern (VoC) and an increasing proportion of vaccinated individuals. This methodology is very innovative in itself, and has not been applied to other datasets before. It involves the application of a new population-based Kalman filter estimate (proposed by the authors in Collin et al. (31)), which extends classical Kalman filters to the parameters estimation across a whole population of subjects. It is to note that association of NPIs with epidemic dynamics has never been formally studied using French data. Finally, it is also noteworthy that placing the estimation of epidemic dynamics into a population approach is singular. More precisely regarding the inference procedure, the first step is to estimate transmission rates in the 12 mainland French regions. This is a major challenge because the available data are very sparse and noisy, and the parametric form of transmission rates is also unknown. By assimilating data across multiple geographic regions and coupling public data with a dynamic mechanistic model, smooth transmission rates can be estimated using a Kalman filter approach (32; 33) – as already used in epidemiology for COVID-19 spread (34; 35; 36) or for other epidemics with regional variability (37). More specifically, we develop a sophisticated method to address this difficult problem based on two important methodological innovations: (1) in the model with the introduction of time-varying dynamics for the transmission rate, including a Wiener process that accounts for modeling errors, and (2) in the way the population is integrated, as we use a new method from the Kalman filter that is compatible with population approaches. This method, in which the log-likelihood function is – estimated by, for example, the unscented Kalman filter (38) – elegantly couples data across multiple geographic regions, is presented in Collin et al. (31). These two innovations are coupled in a strategy that allows estimation of smooth transmission rates without imposing prior-knowledge on their shape or evolution, and relate them to key NPIs, seasonal weather conditions, vaccination coverage, as well as new VoC circulation.

Section Materials and methods presents the supporting data and the underlying mechanistic model for the epidemic, before introducing our two-step strategy to estimate first transmission rates and their associations with NPIs using a multivariate regression approach. Section Results highlights the different effects estimated for the NPIs implemented in France during the first year of the pandemic. Finally the strengths and limitations of our approach are acknowledged in Section Discussion.

## 2 Materials and methods

Open data on the French COVID-19 epidemic, including data on hospitalizations, NPI implementations, VoC prevalence, and the vaccination program are presented below. We then introduce our dynamic modeling for the COVID epidemic, relying on an extended SIR type model. Finally, we describe our strategy for estimating transmission rates using a population-based Kalman filter approach from Collin et al. (31) to study the associations between transmission rates and NPIs, seasonal weather conditions, and VoC.

### 2.1 Available Data

#### 2.1.1 Hospitalization Data

Hospitalization data come from the SI-VIC database (Système d’Information pour le suivi des VICtimes), a government system established in 2016 to identify and track victims in exceptional circumstances (e.g., terrorist attacks). Since beginning of March 2020, the SI-VIC database of the French public health national agency (Santé Publique France) provides the daily number of hospitalized COVID-19 patients, at different geographical levels. In this work, we focus on the 12 non-insular regions of mainland France.

Each entry in the SI-VIC database indicates a patient hospitalized in connection with COVID-19. To qualify, at least one of two criteria must be met by this patient: i) a biologically confirmed diagnosis of COVID-19 (e.g., RT-PCR positive test result) or ii) a chest examination CT scan suggestive of COVID-19. In this analysis, we rely on the daily incident number of hospitalizations (denoted 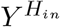) and the total prevalence of persons hospitalized daily (denoted *Y* ^*H*^, corresponding to the number of occupied hospital beds). We consider a period of 391 days (from March 2, 2020 to March 28, 2021). To compare the magnitude of the epidemic in each region, we standardized the data using the population size in each region to present hospitalizations per 100,000 inhabitants. The two data series are displayed in Appendix A, Figure 8.

#### 2.1.2 Non-pharmaceutical interventions (NPIs)

The timing and modalities of the various NPIs implemented in France during the epidemic are taken from the French government’s website summarizing the measures (39). In France, public health interventions were multifaceted. In our analysis, we considered the following summary NPI that took place during the first year of the epidemic in France: i) first lockdown (with two phases of easing/reopening, as described below), ii) second lockdown (with one phase of easing/reopening, as described below), iii) curfew from 8PM, iv) curfew from 6PM, v) closure of schools, vi) closure of bars and restaurants, vii) barrier gestures (including all mandatory hygiene protocols: physical distancing, hand washing, part-time distancing, remote work and wearing masks in public spaces). Of note, NPIs such as travel bans, enhanced testing, contact-trace isolation were ignored to ensure identifiability because they were either of difficult-to-quantify magnitude, or enforced in complete overlap with other NPIs. In addition, partial interventions at the sub-regional level were not taken into account. This resulted in a fairly similar profile of interventions across regions, as most of them were applied simultaneously in the 12 regions of interest. Figure 1 provides an overview of all 10 NPIs considered over time in the *Île-de-France* region. The timing of the introduction of the other NPIs is quite similar in the other regions, differing only by a few days, (1) due to the school vacations (one week postponement) and (2) due to the earlier introduction of the 8pm curfew in the areas most affected by COVID-19, see Figure 9 in Appendix A for more details.

**Figure 1.**
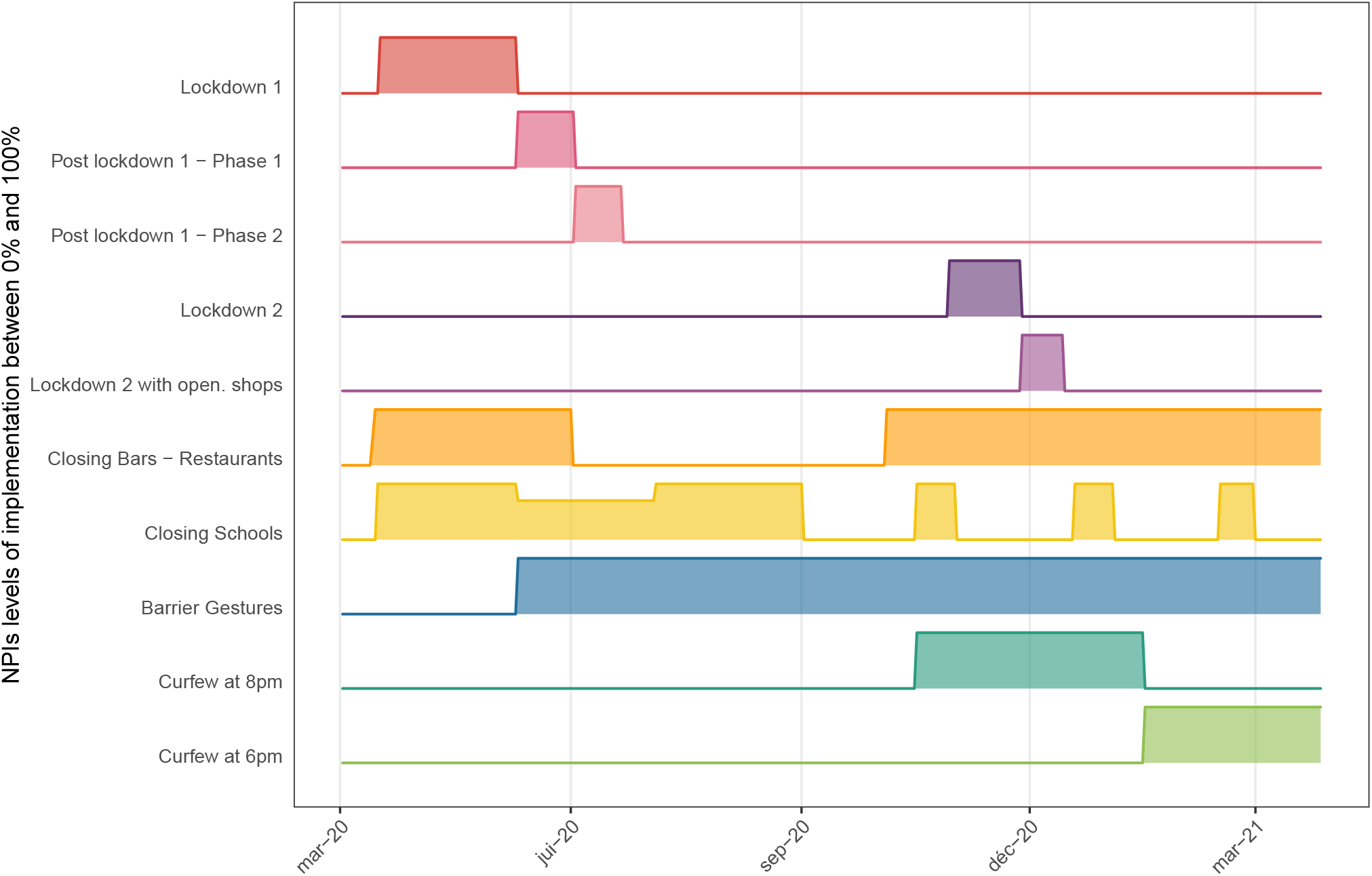
Implementation of major NPIs in Region “Ile de France”. The timing of NPIs implementation is fairly similar across all regions and only differs for a few days in the implementation of school closing because of holidays and curfews 8pm (which was initiated few days earlier in regions with highest COVID-19 burden). All NPI are on/off except school closure right after the first lockdown due to the implemented lock down lifting process as described in the text.

The first and second lockdowns were differentiated because they had distinct modalities resulting in different behaviors and thus potentially impacting transmission differently. For example, during the first lockdown (from March 17, 2020, to May 11, 2020), the entire population was required to work from home - the only exceptions were critical workers from the medical sector, food industry or security, while personal outings could not exceed one hour within a one-kilometer radius from home. During the second lockdown (from October 29, 2020, to December 15, 2020), on-site work was allowed if working from home was not possible, and outings were limited to 3 hours within a 20-kilometer radius. In addition, the end of the initial lockdown was gradually divided by the government into three official phases (Phase 1: May 11, 2020 to June 2, 2020, Phase 2: June 2, 2020 to June 22, 2020, and Phase 3: after June 22, 2020) with many evolving measures such as authorized distance of travel, reopening of cultural sites (*i*.*e*., museums), reopening of non-essential stores, etc. A government campaign to raise awareness of barrier gestures began at the end of the first lockdown. Masks and hand washing were mandatory in many places such as public transportation, schools, and businesses. We thus presume that the *barrier gestures* NPI began on May 11, 2020, assuming that most mandatory measures with potentially high impact have been implemented by that time. Regarding the second lockdown, non-essential stores reopened 2 weeks before the end of the lockdown before Christmas. To account for this, we divide the second lockdown into two phases: a full lockdown until November 28, 2020, and a reduced lockdown thereafter.

School closures were documented by the official holiday schedule, which could vary from region to region. In France, schools were also closed during the first lockdown. In addition, from May 11, 2020 (the end of the first lockdown) to July 4, 2020 (the end of the school year), schools gradually reopened and student enrollment slowly increased back to normal. Estimated student enrollment averaged 30% of school capacity during this transition period according to general media coverage. Full closures of all bars and restaurants occurred twice during the study period: first a few days before the implementation of the first lockdown in March 2020 (as with school closures) and at the beginning of the second lockdown in October 2020. After the end of the first lockdown in June 2020, this measure was gradually lifted in all regions. At the end of the second lockdown, the measure was not lifted and was still in effect at the end of the study period.

Curfew was first enforced on October 17, 2020, in the 12 regions of interest in several large cities and in *Île-de-France* from 9PM to 6AM. It was extended to 54 departments (sub-regional administrative units) on October 22. During the second lockdown from Oct. 30 to Dec. 15, 2020, a national curfew from 8PM to 6AM was en forced. From January 2 to 12, 2021, curfew start at the department level was gradually advanced to 6PM until January 16, when it became 6PM for all the 12 regions of interest. Finally, on March 20, 2021, curfew starting time was changed back to 7PM nationally. For the sake of simplicity, we considered only two different NPIs, grouping together curfews starting at 9PM or 8PM on the one hand, and those starting at 6PM or 7PM on the other hand, and we considered a region under curfew only if it was enforced in the entire region. Assuming that curfews and lockdowns induce different behaviors, curfews are not considered to be included in lockdowns, even though it is forbidden to go out in the evening during lockdowns. Similarly, 6PM curfew and 8PM curfew were considered distinct interventions rather than nested interventions because they are likely to produce different behaviors, and thus have different impact on transmission.

### 2.2 Other exogenous variables: weather, Variants of Concern (VoC), vaccination coverage

#### 2.2.1 Weather conditions

The role of weather conditions in SARS-CoV-2 transmission remains controversial, and early publications have been criticized for their inconsistent results (40). Nonetheless, based on comparisons with other respiratory infections, the potential effects of temperature and humidity on aerosolized and fomite transmission pathways are based on sound mechanistic arguments (41). As the Northern Hemisphere underwent a second winter season during the pandemic, evidence linking transmission to seasonal trends in temperature and humidity (also affecting human behavior and indoor/outdoor gathering) appears more robust (41; 42; 43; 44). Daily weather data – namely temperature in Celsius (*T*), relative humidity in percent (*RH*), and absolute humidity in g.m^−3^ (*AH*) – measured from meteorological stations were extracted from the National Oceanic and Atmospheric Administration database using the R package *worldmet*. All stations located in the region or within 10 km of the region boundary were used to compute daily regional weighted averages (to account for differences in population density within a region, we applied a weighting based on the population within a 10-km radius around each station giving more weight to weather conditions in densely populated areas). We use the Index PREDICT of Transmissibility of COVID-19 (IPTCC), as defined by Roumagnac et al. (45):

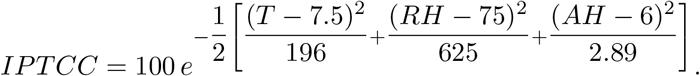

This IPTCC index ranges from 0% to 100%, the smaller the less favorable the conditions for COVID-19 transmission. In France, we observe a seasonal variation of IPTCC low in summer and higher otherwise, with a northeast/southwest gradient. For easier interpretation, we normalized this IPTCC (forcing its range to one), subtracting the national average, and inverting it (see grey curves in Figure 2). Thus, the lower the value, the closer the temperature and humidity conditions were to the optimal transmission conditions defined by Roumagnac et al. (45). Finally, to focus on seasonal variations, a *loess* smoothing with a span of 0.2 was applied, resulting in a smooth weather variable denoted *W* below displayed in Figure 2 for each region. Summer (with higher values of this weather variable) is clearly standing out from winter (with lower values). Taking *W* = 0, we consider the global average value over all French regions during the study period, and *W*_*i*_(*t*) denotes the weather in the *i*^th^ region at date *t*.

**Figure 2.**
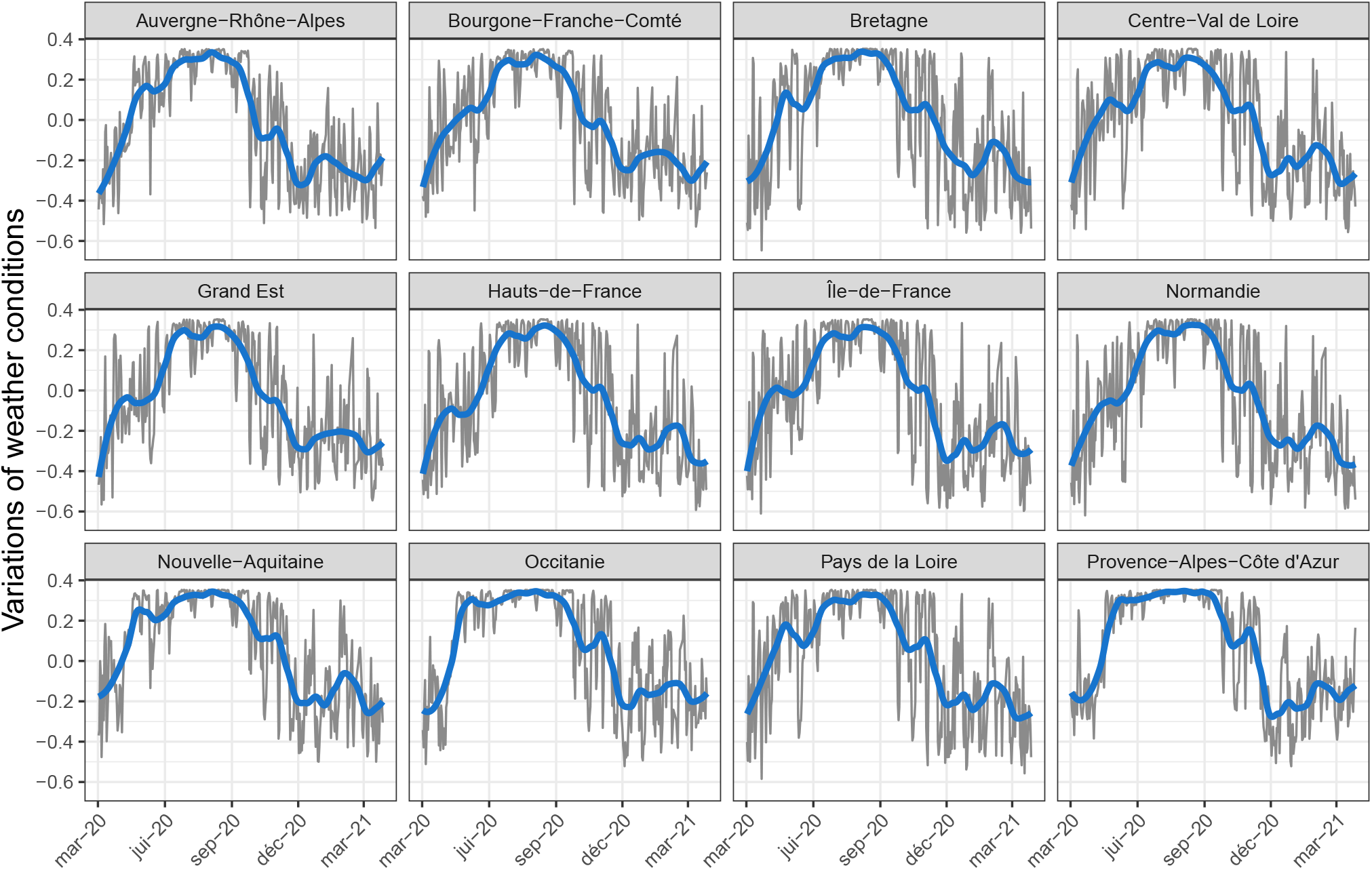
Weather variable modeling the seasonal weather conditions of the 12 regions of interest in grey and after smoothing in blue (denoted *W*). The higher the value, the lower the favorable conditions (temperature and humidity) for transmission of COVID-19.

#### 2.2.2 SARS-CoV-2 Variants of Concern (VoC)

Certain variants of the SARS-CoV-2 virus have been classified as VoC by national and international health authorities because they affect transmissibility or virulence or reduce the effectiveness of interventions (46). Within our study period, French health authorities have conducted surveys to estimate the prevalence of three VoCs: 20I/501Y.V1 (alpha), 20H/501Y.V2 (beta), and 20J/501Y.V3 (gamma). The delta and omicron lineage VoC surveys took place after this study. We therefore included the cumulative proportion of cases infected by any of these three first VoC as a possible covariate associated with transmissiona and we denote this variable as *V oC*_*i*_(*t*) in the following.

We used data from two cross-sectional so-called “flash” surveys conducted on January 7 & 8, 2021, and on January 27, 2021 (47; 48), as well as the weekly estimate of VoC spread provided by the SI-DEP database at the regional level from February 12, 2021, to March 28, 2021 (49). Between January 8, 2021, and February 12, 2021, the estimated proportion of the sum of the three VoC increased from a national average of 3.3% to a national average of 46%. To fill in the missing data, we assume that the proportion is equal 0% before January 8, 2021, and that the trend is linear between January 8 and January 27, 2021, and linear between January 27 and February 12, 2021. It should be noted that a logistic and an exponential growth were also investigated, without resulting in any significant change in our conclusions (results not shown). Since no data were reported for the *Bourgogne-Franche-Comté* region on January 27, 2021, only one slope was estimated from January 8 to February 12. VoC fraction in each region is displayed over time in Appendix A.

#### 2.2.3 Early vaccination

Vaccination began in France on December 27, 2020, at which time three COVID vaccines were licensed and available: BNT162b2 mRNA (Pfizer), ChAdOx1 nCoV-19 (AztraZeneca), and mRNA-1273 (Moderna). To account for the starting vaccination campaign, we used the database VAC-SI (50), which records the cumulative percentage of the population vaccinated with at least one dose of vaccine over time. Vaccination was initially prioritized for the elderly aged 75 years and older. The proportion of the total population vaccinated increased to approximately 12% by the end of the study period, and regional population vaccination coverage rates over time are shown in Appendix A.

### 2.3 Modelling the epidemic

#### 2.3.1 The mechanistic model

We model the evolution of the COVID-19 epidemic using an extended SEIR model (51; 52), called the SEIRAH model, in which the population of size *N* is divided into 5 compartments: susceptible *S*, latently exposed *E*, symptomatically infectious *I*, asymptomatic/pauci-symptomatically infectious *A*, hospitalized *H*, removed *R* (i.e., both recovered and deceased), see Figure 3. The number of vaccinated individuals denoted by *V*, is assumed to be known, see Section 2.2.3. The dynamics of such a model is given by

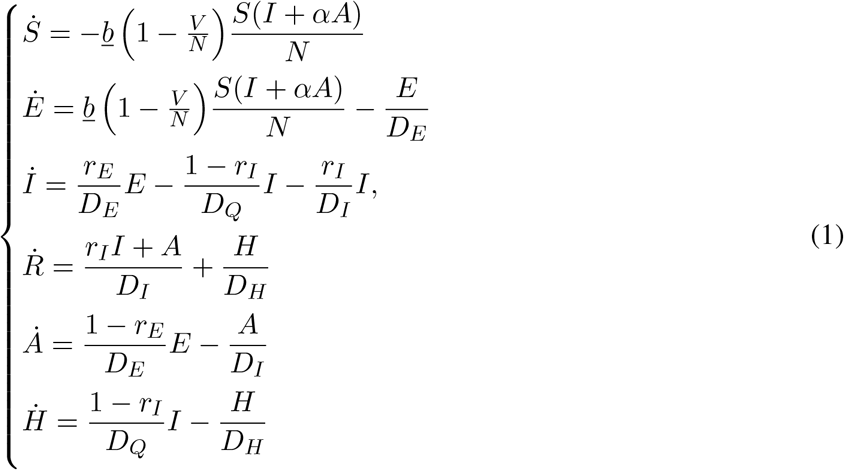

where *α, r*_*E*_, *D*_*E*_, *r*_*I*_, *D*_*I*_, *D*_*Q*_, *D*_*H*_ are time-independent parameters described in Table 1 while *b* is a function of time modeling the disease transmission rate.

**Figure 3.**
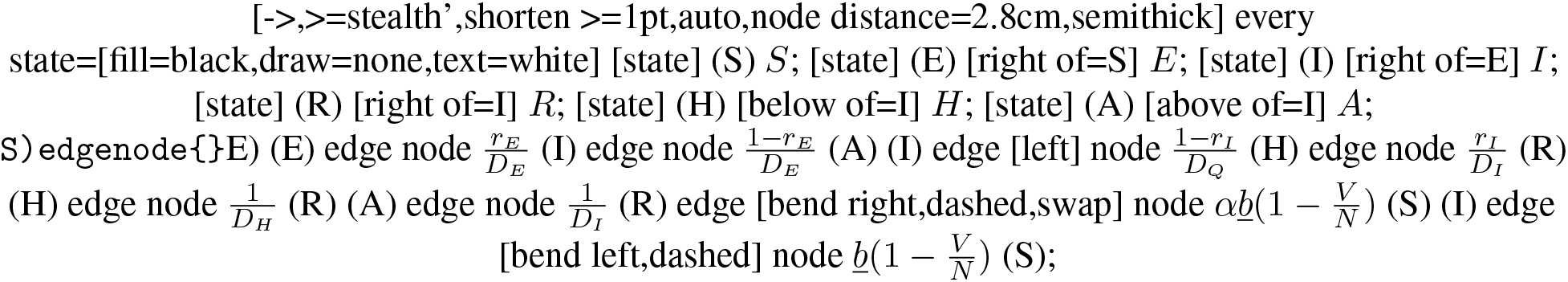
SEIRAH model representation

**Table 1.**
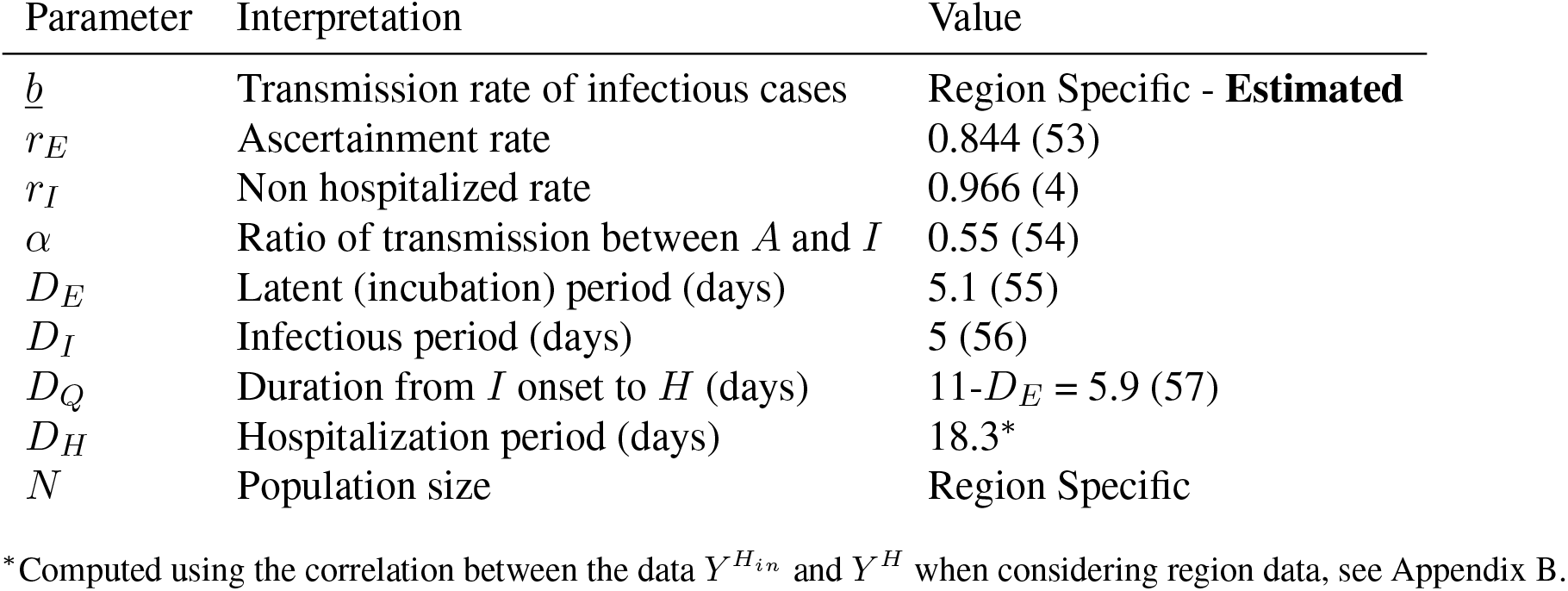
Model parameters for the SEIRAH model and associated values.

| Parameter | Interpretation | Value |
| --- | --- | --- |
| $\underline{b}$ | Transmission rate of infectious cases | Region Specific - **Estimated** |
| $r_E$ | Ascertainment rate | 0.844 (53) |
| $r_I$ | Non hospitalized rate | 0.966 (4) |
| $\alpha$ | Ratio of transmission between $A$ and $I$ | 0.55 (54) |
| $D_E$ | Latent (incubation) period (days) | 5.1 (55) |
| $D_I$ | Infectious period (days) | 5 (56) |
| $D_Q$ | Duration from $I$ onset to $H$ (days) | $11 - D_E = 5.9$ (57) |
| $D_H$ | Hospitalization period (days) | 18.3* |
| $N$ | Population size | Region Specific |

\*Computed using the correlation between the data $Y^{H_{in}}$ and $Y^H$ when considering region data, see Appendix B.

#### 2.3.2 The observation model

The two quantities *Y* ^*H*^ and 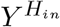 relate to the solutions of System (1) respectively as for all regions *i* = 1, …, 12, for all observation time in days *j* = 1; … ; 391: 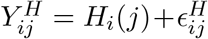 and 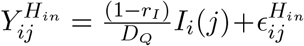, in which 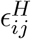 and 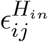 represent normally distributed constant measurement errors.

#### 2.3.3 Effective reproductive number and attack rates

When individuals are homogeneous and mix uniformly, the effective reproductive ratio *R*_eff_(*t*) is defined as the mean number of infections generated during the infection period of a single infectious case at time *t*. In this model, the effective reproductive ratio can be written as a function of model parameters (see Appendix C for details):

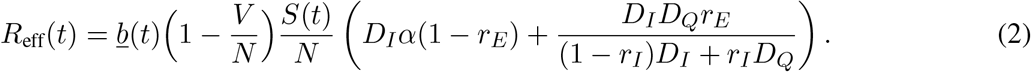

When neglecting the deaths, the proportion of infected individuals assuming no waning immunity – also called attack rates – among the population in each region at a given date is given by:

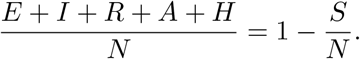

### 2.4 A population-based Kalman filter to estimate the transmission rate

#### 2.4.1 Framing the problem

To support the modeling decisions underlying the disease transmission rate *b*, and more generally the inference procedure chosen, we applied Kalman-based estimation strategies (32; 33) to the epidemic model (1). We define the global transmission rate *t* ↦ *b*(*t*), which accounts for the proportion of susceptibles removed from the system by vaccination:

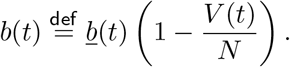

We propose to estimate *b* and finally *b* using an estimate of vaccinated individuals *V* (see Subsection 2.2.3). In each region *i*, we then introduce a dynamic equation for *b* of the form

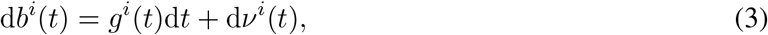

where *ν*^*i*^ consists of a Wiener process such that for all *t, s* ≥ 0, *ν*^*i*^(*t*) − *ν*^*i*^(*s*) ∼ 𝒩 (0, (*t* − *s*)*σ*_*ν*_) with 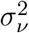 known and constant across all regions, and *g*^*i*^ a function describing the evolution of global transmission rates *b*^*i*^ in each region.

After discretization using Euler’s time scheme with a sufficiently small time step *δt*, we obtain a discrete-time dynamical system applied to the variable *x* = (*E, I, R, A, H*) ⊤ ∈ ℝ^5^, for each region *i* = 1, …, 12:

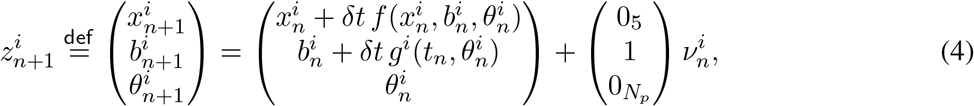

where *f* accounts for the dynamics of (*E, I, R, A, H*) in (1), while *S* is subsequently reconstructed as *S* = *N* − (*E* + *I* + *R* + *A* + *H*) in each region. In the discrete-time system, 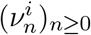 now represent independent random variables, normally distributed with 0 mean and variance equal to 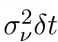. Moreover, if constant parameters need to be estimated, the vector 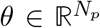 summarises all these parameters. For the estimation below, we transform the variable *z* to account for biological constraints. Since all state variables are positive and bounded by *N*, the total population size of the region, we transform the state variable with *x* ↦ logit(*x/N*). We also apply a similar transformation to *b* ↦ logit(*b/*max_*b*_). Note that the state and transmission rate variables in the transform space tend to have Gaussian distributions. After calibration, we set max_*b*_ = 1.5 and verified that other values did not significantly change the results (result not shown).

#### 2.4.2 Population approach

To perform the estimation, we rely on an extension of the classical Unscented Kalman Filter (UKF) (58; 38; 32). The peculiarity of this application is that multiple data series in multiple regions are observed together, since we observe multiple realizations of the same epidemic (each region being a different realization). To account for parameter correlation across the different regions in our Kalman estimation, we follow a recently proposed population-based Kalman formulation (31). As in mixed effects models (59), each initial uncertainty variable 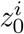 is assumed to be randomly distributed around a common population intercept 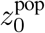 with a Gaussian distribution of unknown covariance ***Q***_0_, namely:

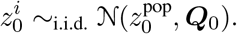

By treating the population intercept as an empirical mean over the population members, we obtain a classical filtering problem (32) on the aggregate variable 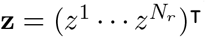 when constructing our objective function. The only difference is the formulation of the initial covariance prior 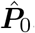, which couples observations across regions and can be written as follows:

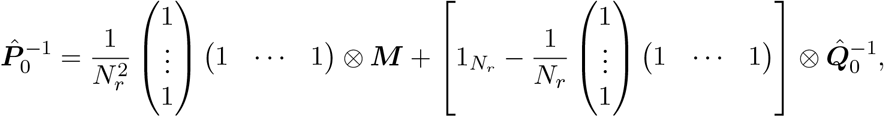

where ⊗ denotes a Kronecker product,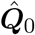 is a prior of ***Q***_0_, and ***M*** is a small penalization matrix guaranteeing the overall invertibility of 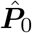. As a result, the matrix 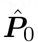 is non-block diagonal with respect to the region *i* and thus all the dynamics of the regions are coupled. The resulting discrete-time Kalman estimator ties all regions together to obtain a population-based estimate. Note that in such a strategy, it is possible to force a variable in the population to be constant by simply choosing 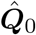 such that 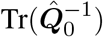 is very small with respect to Tr(***M***). Conversely, a small Tr(***M***) with respect to 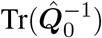 in a large population *N*_*r*_ ≫ 1 will encourage regions to remain independent from each other. Given prior knowledge, our Kalman implementation uses the available measurements 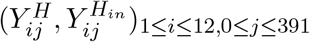 to recursively compute the following estimates:

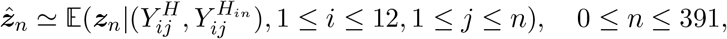

and

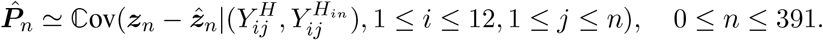

Taking advantage of 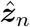 gathering the augmented state 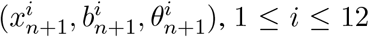 of the 12 regions, a simple post-processing over the regions yields estimates of *b* and the state variables. The fact that the state variables S, E, I, R, A, and H are included in the augmented state means that the errors in the initial conditions are corrected over time, allowing for different prevalences in different regions. We refer the reader to Collin et al. (31) for more details on this Kalman-based population approach.

#### 2.4.3 Estimation strategy

Since we want to inject as little information as possible about the shape of the transmission rate *b*, we will first assume that the Wiener process *ν* defined in Equation (3) is a time-dependent function, thus encompassing the entire dynamics of *b*. This is consistent with setting *g* ≡ 0 (indicating no prior knowledge of the relationships between the evolution of the transmission rates and the NPIs). However, to avoid overfitting, our goal is to distinguish the latent trajectory of *b* from noise in the observations. We use a 3-step approach for smoothing the trajectories of *b*, described below:

1. Estimate an appropriate prior for the initial transmission rates *b* before the start of any NPI. We use data before the first lockdown (10 days available) and assume that the transmission rate 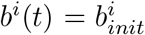 is a constant for *t* = 1 … 10 days. In other words: We apply the population Kalman filter estimate described above, with *θ* from Equation (4) reduced to *b*_*init*_.
2. Estimate the shape of *b*(*t*) with a prior for the initial value, but without a prior for the dynamics. We set the initial value of *b*^*i*^ for times *t* = 1 … 10 days to 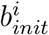 and take *g*(*t*) ≡ 0 such that Equation (3) rewrites d*b*(*t*) = d*ν*(*t*). We apply the population Kalman filter estimation described above. Note that the parameter vector *θ* from Equation (4) is now empty, and since there is no information about *b*, the model error is very large and could lead to overfitting. We then create a prior for the dynamics of *b* by fitting a parametric form based on the sum of logistic functions (well suited to modeling observed variations such as stiff lockdowns or smooth unlocks) to the weighted average trajectories of *b* over all regions using the least squares method. The number of logistic functions summed is based on the observed number of principal changes in the variations of the weighted average trajectories of *b*. This now gives us a prior *g* for the dynamics.
3. Estimate the shape of *b* with a prior for the initial value and an informative prior for the dynamics. Finally we set the initial value of 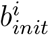 for times *t* = 1 … 10 days to *b*^*i*^ and take d*b*(*t*) = *g*(*t*)d*t* + d*ν*(*t*) as in Equation (3). We apply the population Kalman filter estimation described above. Since the dynamics of *b* still contains modeling noise, the shape of *b* differs from the prior transmission rate defined in Step 2. This final estimate will be used to further describe the relationships between transmission rates and NPIs.

### 2.5 Explanatory model for the transmission rate

#### 2.5.1 Mixed effects model

Using the *b* estimate obtained from the population-based Kalman filter, in the third step described above, we can determine the association between NPIs, seasonal weather conditions, VoC proportion and the transmission rate 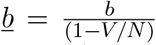. We followed Flaxman et al. (19) and considered interventions to have multiplicative effects. Therefore, we applied a linear mixed effects model to the log transformation of transmission. The model equations are given in Appendix F. It consists of an intercept (representing the average transmission of COVID-19 over all regions without NPIs, without VoC circulation, and for average French weather conditions (*W* = 0)), the effects of the 10 NPI described in Section 2.1.2, the effect of weather, and the effect of VoC fraction. Because the transmission of SARS-CoV-2 is different indoors and outdoors (60), we also added an interaction effect between bar and restaurant closure and weather that accounts for the opening of outdoor seating areas. This is the only interaction included (and studied) to avoid overfitting. Finally, we added random effects to account for heterogeneity between regions. We added a random intercept and random slopes for the effect of the first and second lockdowns as well as the 6PM curfew because their effects can vary greatly between regions. We assume a full covariance matrix for the random effects, so that the associations in each region may be correlated – in particular, they could be influenced by several factors not accounted for in the epidemic model, such as population density, age distribution, or urbanization.

Note that we have delayed the lockdowns by 7 days. This can be interpreted as a necessary period of time to allow people to organize and adapt (introduction of home-based work, child care, etc.). This decision was motivated by the observed 7-day delay in immediate transmission rates decline obtained with Kalman filters, see 4. To facilitate interpretation of the estimated associations, the parameters were transformed and expressed as percent decrease or increase in transmission by applying the function *x* ↦ 100(*e*^*x*^ − 1). Classical 95% confidence intervals were obtained as 100(*e*^*x±*1.96*SE*(*x*)^ − 1) assuming normality, where *SE* is the standard error obtained from the regression.

#### 2.5.2 Interpretation of the association between seasonal weather conditions and transmission rates

The variable for seasonal weather conditions being unitless, so interpretation of its estimated effect is done in comparison to the average weather conditions in France. To further facilitate understanding, we computed the average relationships between NPIs and transmission rates during both summer and winter periods respectively, using the summer (or winter) average of *W* (*t*) from June 21^st^, 2020 to October 21^st^, 2020 (or before June 21^st^, 2020 and after October 21^st^, 2020) across all regions.

#### 2.5.3 Basic reproductive number

The intercept of the above regression represents the mean transmission rate over all regions when there is no NPI and no VoC and the weather conditions are assumed to be the average weather conditions over a year in France. Thus, substituting it into Equation (2), directly provides the basic reproductive number and 95% confidence intervals can be calculated using the standard error of these parameters.

## 3 Results

### 3.1 Estimation of the transmission rate using a population-based Kalman filter

Step 1 of the estimation provided the initial values for the transmission rate with fairly similar values across regions (average 0.78 sd 0.012, see details in Appendix D). Although higher values were found in regions where the first wave was stronger, the low variability between regions suggests that the magnitude of the first wave is not solely due to a higher transmission rate, but also to differences in epidemic baseline conditions (i.e., the number of exposed and infectious cases, linked to differences in the timing of virus introduction or the occurrence of super-spreading events (61)) and weather conditions. Transmission rates estimated in Step 2 without knowledge of their shape are shown in Figure 4 (top, left) for all regions. Since there is no prior information about *b* inputted, the model error is very important and leads to over-fitting of the data. In particular, many regions exhibits weekly oscillations related to the under-reporting during weekends. We approximated the weighted average trajectory of *b* by a sum of 7 logistic functions is, as shown in Figure 4 (bottom, left) which becomes our prior in Step 3. Finally, in Step 3, smooth regional transmission rates *b*_*i*_ are estimated, see Figure 4 (top, right). Since the dynamics still contain modeling noise, the shapes of regional *b*_*i*_ are naturally different from the prior defined in Step 2 (while still smooth).

**Figure 4.**
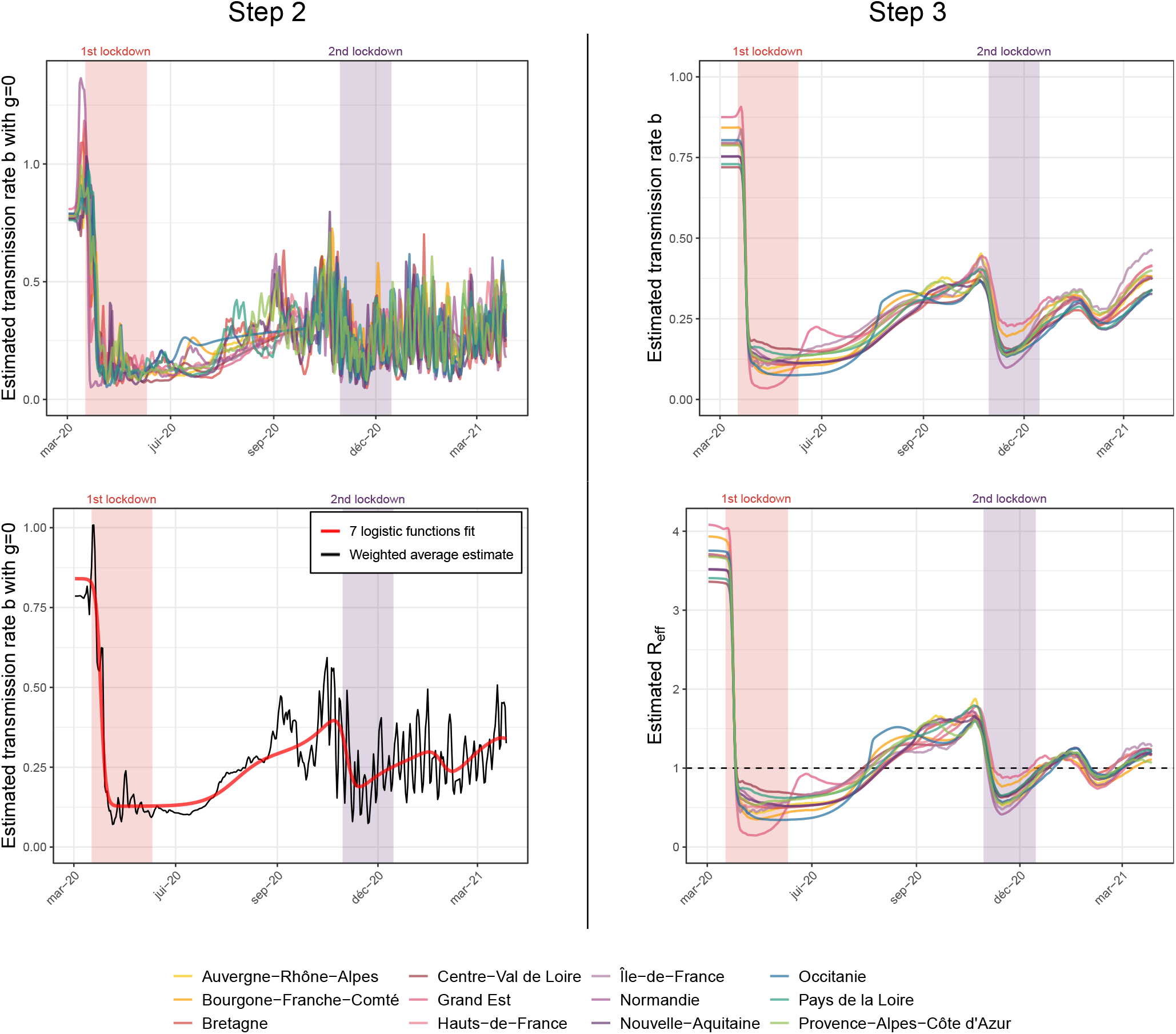
First column (Step 2) Top: regional dynamics of *b* with *g* = 0 (without *a priori*). Bottom: mean value of *b* over time (black line) obtained in Step 2, fitted with 7 logistic functions (dashed red line). **Second column (Step 3)** Top: estimated regional dynamics evolution of *b* with *g* the 7 logistic functions obtained in Step 2 (with *ν* ≠ 0). Bottom: estimated regional dynamics of *R*_eff_ with *g* the 7 logistic functions).

### 3.2 Basic and Effective reproductive number

Figure 4 (bottom, right) presents the estimated effective reproductive ratio *R*_eff_, with starting values ranging between 3.5 and 4 in all regions. This variability could be due in part to winter weather conditions in early March 2020, and after adjusting for weather condition we estimate the national average basic reproductive number at 3.10 [2.95 ; 3.26]. Variations of *R*_eff_ over time shows that it quickly falls below the critical value of 1 after initiation of the first and second lockdowns.

### 3.3 Attack rates

The attack rate (defined in Section 2.3.3) provides additional knowledge about the number of possible hidden/unmeasured cases. Figure 5 displays the attack rate at several key points in time: at the end of the first lockdown (May 11, 2020), on October 5, 2020, and at the end of our study period (March 28, 2021), while we estimate national attack rates at these time points to be 5.7%, 8.8%, and 25.3%, respectively.

**Figure 5.**
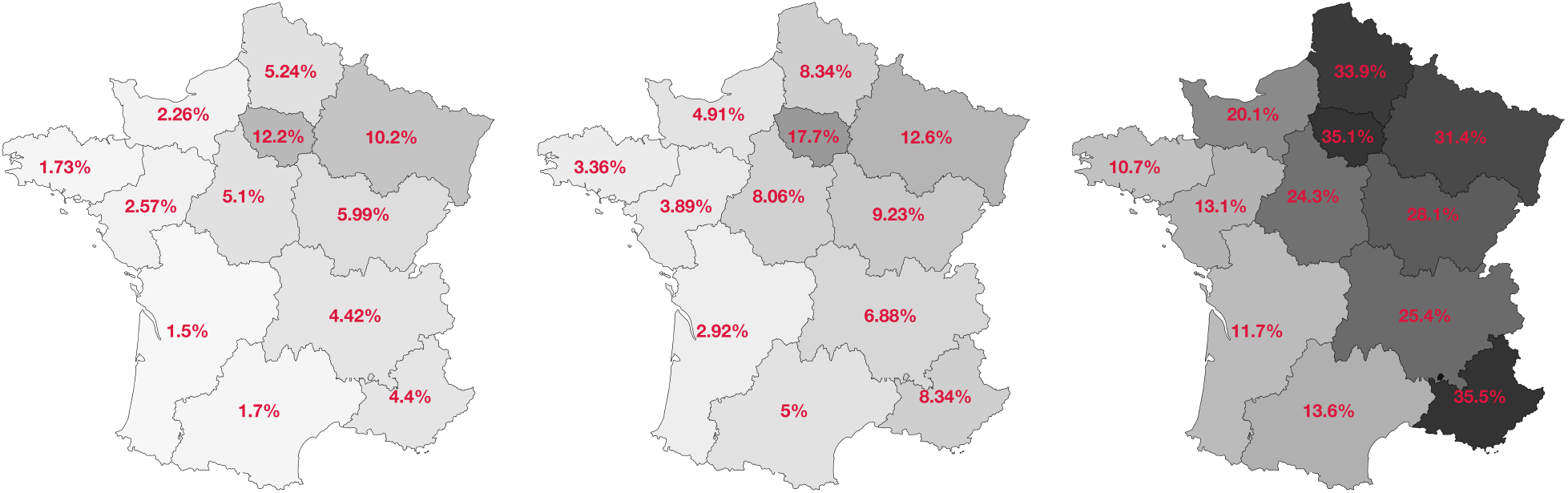
Model estimation for the proportion of naturally immunized individuals in the population (deaths and vaccinated people not taken into account) on May 11^th^, 2020 (left), on Oct. 5^th^, 2020 (middle) and on March 28^th^, 2021 (right).

### 3.4 Associations between NPIs and transmission rates

Table 2 summarizes estimated associations between the NPIs and the transmission rate (from the model introduced in Section 2.5 – residuals and detailed model coefficients can be found in Appendix F). Figure 6 shows the corresponding regional fits. Of note, the values for both curfews are very close (near 30%), and not statistically different (estimated difference is 3% [-2% ; 8%]). We show that all NPIs in this analysis reduce transmission and have an effect that is significantly different from zero.

**Table 2.**
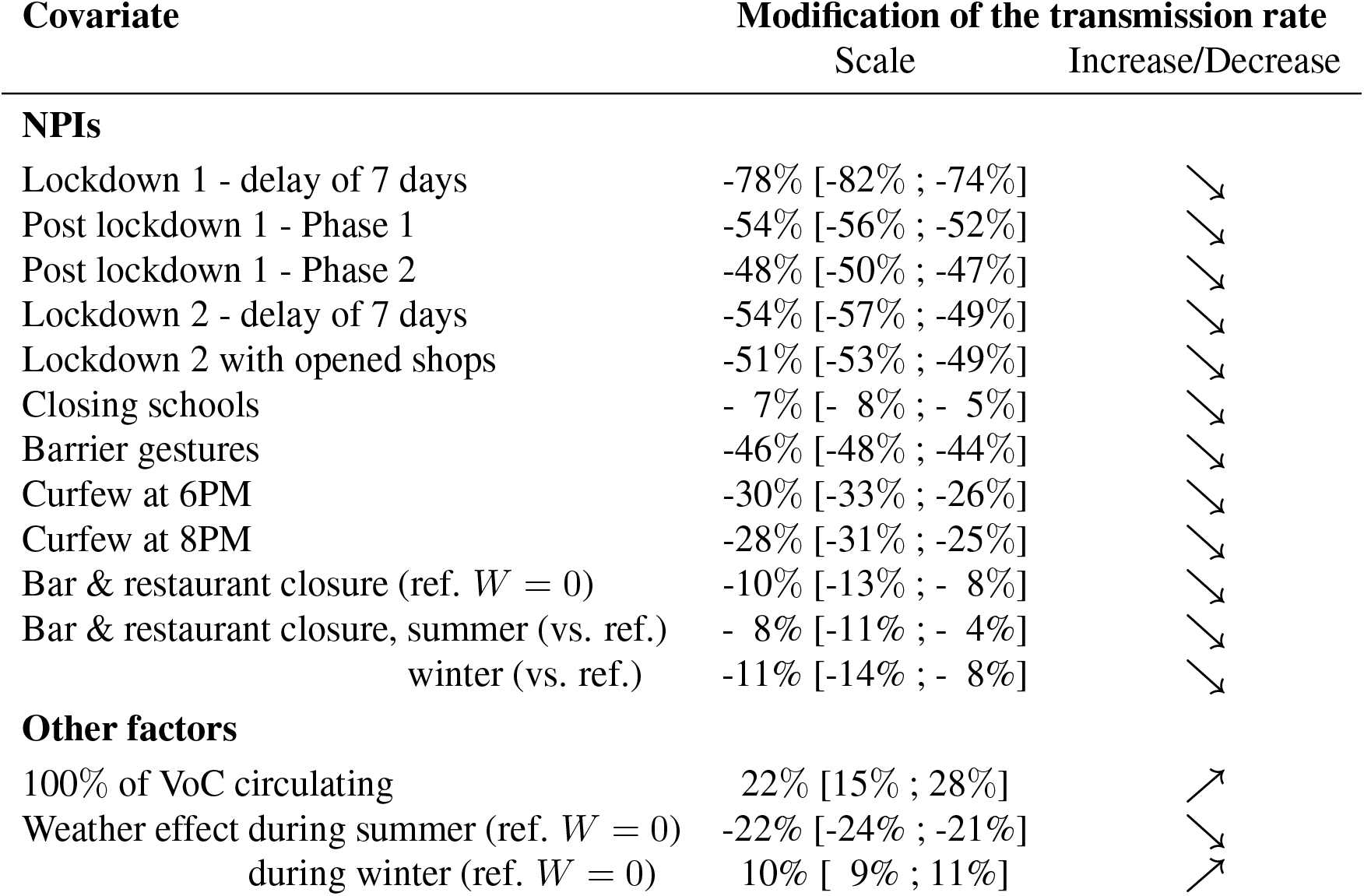
Estimation and 95% confidence intervals of the associations between the transmission rate and seasonal weather conditions, VoC proportion, and NPIs. Model AIC = -1,388.

**Figure 6.**
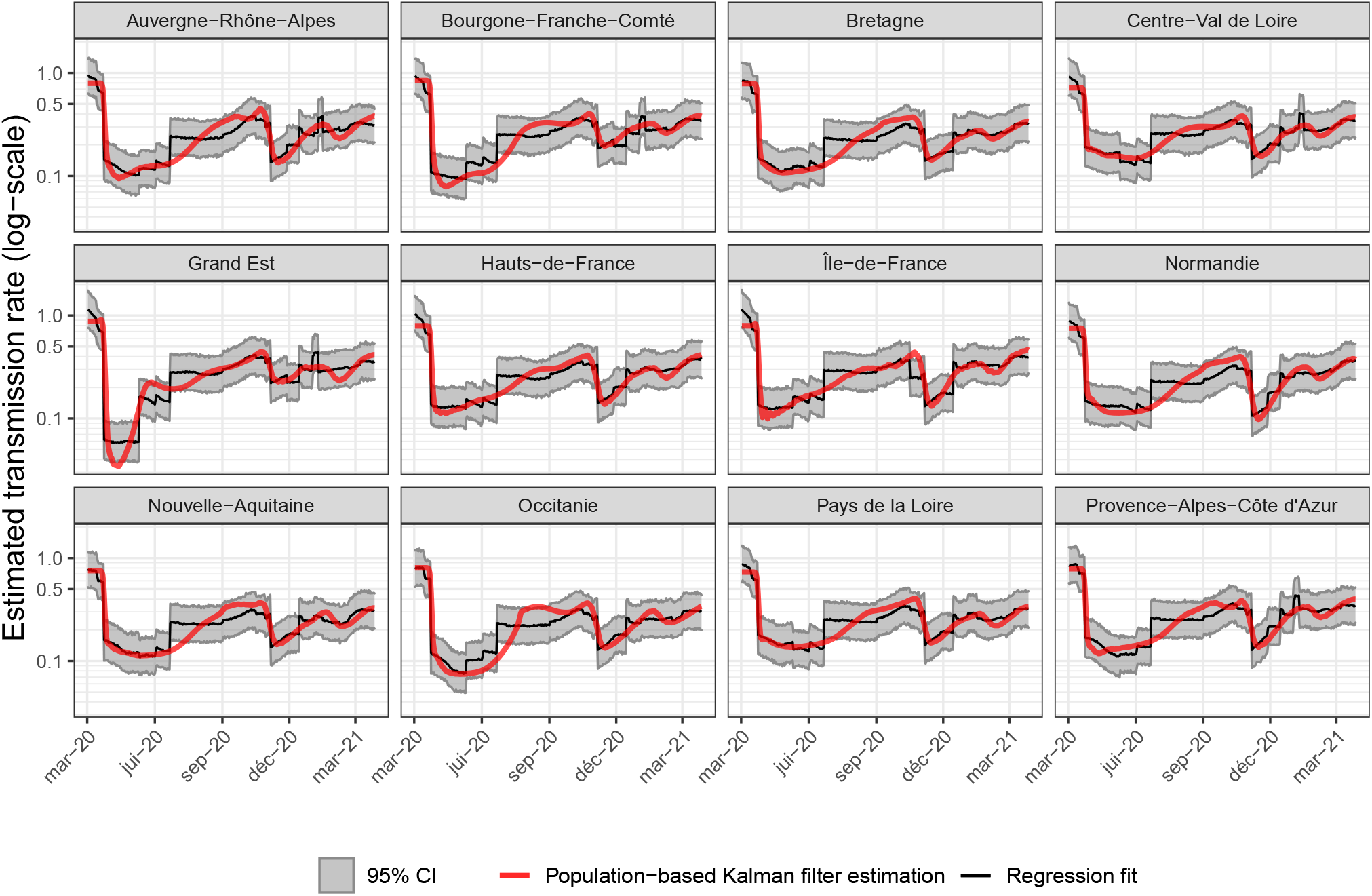
Regression fit of transmission rates from the population-based Kalman filter for each region with random effects on log(*b*_*init*_), *b*_*l*1_, *b*_*l*2_ and *b*_*curf*6*P M*_.

### 3.5 Effect of weather

We estimate that, on average, transmission is significantly increased by 10% during the winter period and significantly decreased by 22% during the summer period (compared to the transmission under average weather conditions). Figure 7 (top) shows the estimated impact of seasonal weather conditions on transmission during the study period in each region. The estimated interaction between bar and restaurant closures and seasonal weather conditions is statistically significant (p=0.037), and complex to interpret. We show that while bar and restaurant closure always has a significant effect and reduces transmission, it is somewhat more effective in winter (11% [8%;14%] decrease) than in summer (8% [4%;11%] decrease).

**Figure 7.**
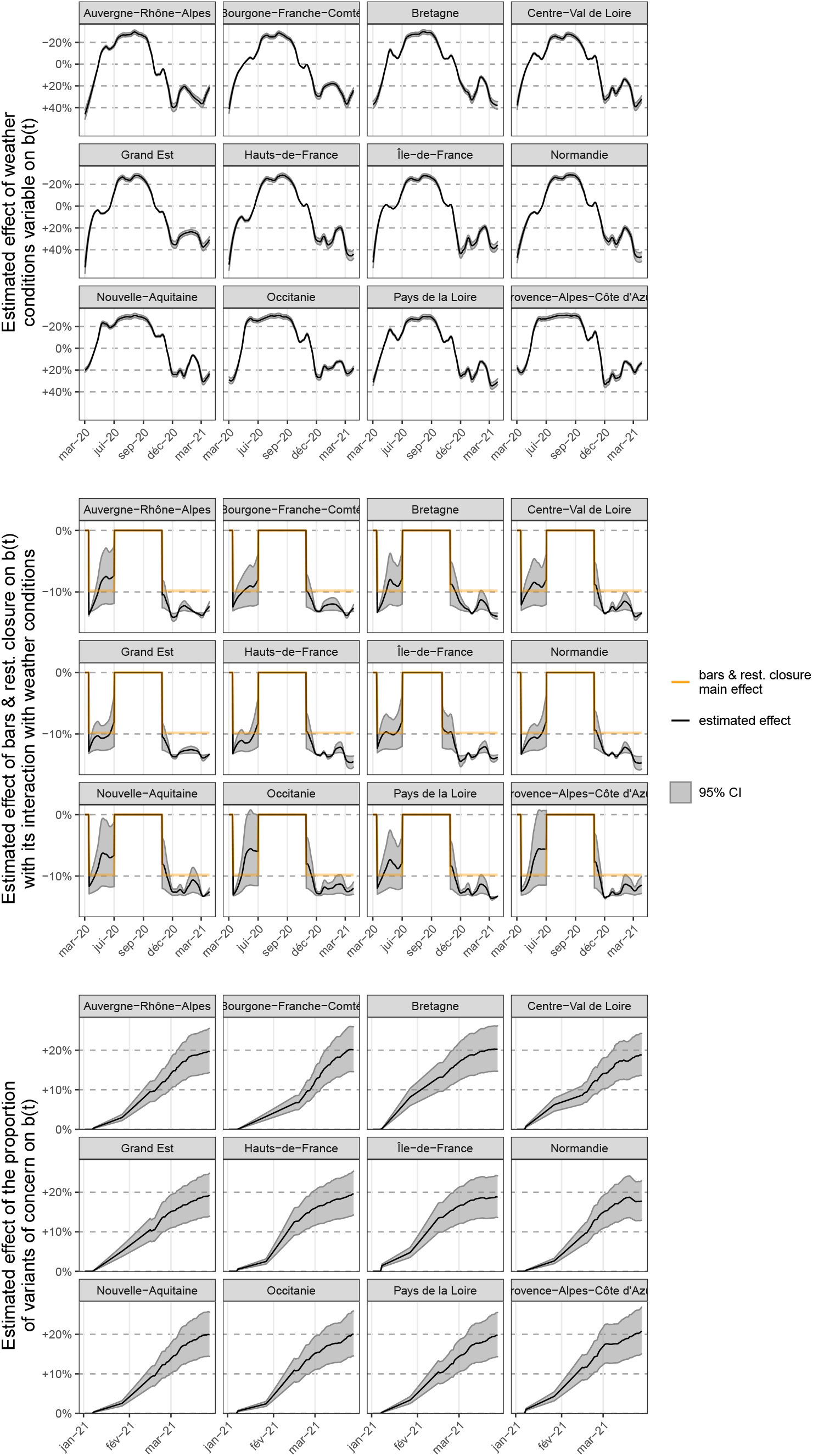
Top: Estimated association between transmission rates and the seasonal weather conditions and their 95% confidence interval for the 12 regions of interest during the study period using the global average during the period as a reference for comparison. Middle: Estimated association between transmission rates and bar and restaurant closures for the 12 regions of interest during the study period. In red, the main effect of -10%. In black and grey, the effect with the interaction with weather conditions and its 95% confidence band. Bottom: Estimated association between transmission rates and VoC occurrence and its 95% confidence interval from January 1, 2021.

Figure 7 (middle) shows the estimated effect of bar and restaurant closures for the 12 regions of interest during the study period and their interaction with weather conditions. Coherently, the impact of weather conditions yields a stronger effect of bar and restaurant closures on reducing transmission in northern regions compared with southern regions (that can be interpreted as the closure being more effective in northern regions where weather conditions are also more favorable to transmission). In addition, a stronger effect is observed in spring 2021 due to weather more favorable to transmission compared to the previous year.

### 3.6 Effect of VoC

Regarding the effect of the three VoC, our results show that increasing the proportion from 0 to 100% would increase the transmission rate by an estimated 22% [15% ; 28%]. Figure 7 (bottom) shows how the increased transmissibility varies over time due to the VoC fraction increasing to nearly 100%.

## 4 Discussion

We propose an innovative method to infer transmission rate over time from hospital data and estimate the association between transmission rates and multiple NPI, weather, and VoC. We show that all considered NPIs have a statistically significant and independent effect on transmission rates. In addition, we demonstrate a strong effect of weather conditions, decreasing transmission rate in summer and increasing it in winter, and we also observe increased transmissibility associated with VoC proportion increase.

Interpretation of our results on transmission rates is conditional on the mechanistic model presented in Figure 3 as well as in the parameter values set from the scientific literature and listed in Table 1. This model does not take into account the age structure of the population. However, we believe that using a population-based approach and relying on random effects takes into account several intrinsic characteristics – such as population density, age structure, transport habits, etc. – that influence the spread of disease through region-specific transmission rates. Another shortcoming of the SEIRAH model is that it does not account deterministically for travel between regions. However, since the state variables S, E, I, R, A, H are also corrected over time using the Kalman filters, our strategy can account for, for example, decreases or increases in the number of exposed individuals E in a region. This means that even if we do not directly model the interaction between regions, we are able to account for the most important changes that could be due to travel between regions. It could be further improved without changing the Kalman methodology by adding a modeling error operator applied specifically to this variable, provided we are able to quantify the degree of fluctuation in the population of each region due to travel. To do more, we could modify the model to include travel between regions, but this would require us to specify additional parameters that are difficult to calibrate without more data on travel between regions. Because the study period was short, vaccination was just starting over the end of the study period and immune escaping VoC were not circulating yet, waning immunity is also not taken into account. Regarding the study data, it is well known that there may be some misreporting in the SI-VIC database, especially when patients are transferred between hospitals or when the number of admissions is under reported during the summer months. However, one might expect these discrepancies to be less significant than for other data sources such as the daily number of new confirmed cases, where testing procedures (and availability) have changed significantly during the epidemic, or for primary care visits, where clinical diagnosis may be less accurate.

Interestingly, although the models were different and the data were not completely identical, our results were comparable in terms of attack rates to existing modeling work (62) and seroprevalence sampling studies (63; 64; 65) (for a comparison, see Appendix E). Overall, our estimates tend to be slightly higher, within a 5% margin. This is probably due to the strong assumption that there is no waning immunity in our model. Indeed, we assume that all individuals, once infected, have a sufficiently high antibody titers to systematically test positive in seroprevalence studies. Regarding the reproductive numbers, we estimate the average national basic reproductive number to be 3.10 [2.95 ; 3.26]. This differs from the initial values of *R*_eff_ in mid-March in all regions, which are estimated between 3.5 and 4, and is mainly due to winter weather conditions in mid-March in all regions. Our estimate is consistent with other estimates worldwide. In Liu et al. (77), the authors compare 12 studies that estimate the basic reproductive number for COVID-19 from China and overseas. Estimates ranged from 1.4 to 6.49, with a mean of 3.28. French studies also found similar estimates 3.18 [3.09 ; 3.24] in Di Domenico et al. (30) and 2.90 [2.81 ; 3.01] in Salje et al. (3). These comparisons validate our estimation strategy, which has the major advantage of estimating transmission rates without making any assumptions about the shape of its dynamics over time. Moreover, the computation times are very reasonable (a few minutes for the full estimation on a regular laptop without code optimization).

Other variables could have been added, such as partial interventions (e.g., only in large cities in regions heavily affected by COVID -19) - but these would have been inconsistent with our model defined at the regional level. Indeed, we need a match between hospital data-used to estimate transmission rates - and NPIs - used to estimate their associations with transmission rates - to combine the two steps of our study. To better illustrate, even if we have more information about bar closures in a city, we do not know exactly what proportion of that city’s population is included in the hospital data for that city’s region. And each region may have a different number of major cities, each of which has a different size in terms of population. So the impact of “partial bar closure” could vary from region to region, which is not consistent with the assumption of the regression.

Other potential variables were grouped under the NPI “barrier gestures” because they suffer from inconsistent definitions and are sometimes allocated simultaneously, leading to identifiability problems. Work from home is a good example. First, compliance with this measure (which can vary due to employee fatigue, organizational difficulties, and lack of legal requirements) is difficult to assess. Second, in terms of quantitative indicators, DARES has conducted surveys of companies with 10 or more employees and has shown that the average percentage of working from home is 30% [19%, 40%] and peaks during the two lockdowns (66). However, this indicator takes into account work from home and paid leave, which can be very different in terms of risk behavior and overlap to some extent with school closure. Gathering restrictions is also a good example of how policies were constantly changing between private and public measures with the size of authorized gathering also varying. Finally, we believe that an individual’s overall compliance with an entire package of “barrier gestures” is more likely to be stable over time than compliance with the individual components of that package. Mask wearing is a good example of this, as people were encouraged to keep the mask even though there was no longer an obligation, and probably did even more when at risk of infecting others. All in all, removing work from home, gathering restrictions, social distancing, mask wearing, and others from the regression model captures their association with transmission rates through the effect of lockdowns and barrier gestures in a more identifiable manner leading to more robust results.

We also make certain modeling assumptions. For some interventions, such as lockdowns, we considered a 7-day delay in implementation. This choice is not determined by the delay between infection and hospitalization, which is already accounted for by *D*_*E*_ and *D*_*Q*_, but by the transmission rates obtained with the Kalman filter. This choice is supported by other studies, such as Dehning et al. (24), in which the delay can be as long as 15 days. However, we examined how this affects our regression fit. Not accounting for the 7-day lag significantly degrades the fits, see Model 4 of Appendix G in the Supporting Information for more details. Modeling the impact of partially opening schools during the period from May 11 to July 4, 2020, as 70% of the impact of a full closure might be considered an oversimplification. During this period after the first lockdown, schools reopened very gradually in three distinct phases, and enrollment increased even more progressively. By June 2^nd^, there were sharp differences between regions in opening and school attendance, with an average of only 30% of students under 12 attending school. The other levels of secondary schools (“collèges” and “lycées”) reopened in early June and gradually enrolled more students until the vacations, which began in early July. To avoid risking identifiability problems, we choose not to differentiate by region or phase of reopening, and a ratio of 0.7 was used for the proportion of closure for all regions. We found modest but significant effect of school closure as of other studies (76). Regarding the effect of curfews, we do not detect a statistically significant difference between 6PM and 8PM. This could be due to an identification problem, as the 8PM curfew was in place for less than 3 weeks in many regions. An alternative explanation could be that both curfews affect global social gathering in the same way. For example, both prevent most private dinners and parties (or at least drastically reduce the number of guests).

We consider weather conditions relying on an aggregated indicator instead of exploring the impact of temperature, absolute humidity, and relative humidity separately. In Appendix G, simpler models are examined: all of them exhibits degraded fits. However, we remain extremely cautious in interpreting the estimated association of weather conditions with transmission rates and the mechanisms potentially involved. Finally, we consider an interaction between seasonal weather conditions and the closure of bars and restaurants, which is attributed to the use of patios and terraces (which have been expanded in many places since the beginning of the pandemic). Of course, other interactions and more complex models can be considered, but this quickly leads to overfitting.

During our study period, the 20I/501Y.V1 (alpha) variant seems to have always been predominant (over 90% at any time) among VoC (the other two being beta and gamma) considered in mainland France (49). Very different estimates of the increase in transmissibility for the 20I/501Y.V1 (alpha) variant have been proposed in the scientific literature, ranging from 29% to 90% (67; 68; 69; 70; 71), with the lower values being consistent with our own results. The higher estimates could be explained in part by the fact that new VoC appeared early in the winter in England and the United States. This could lead to confusion bias between weather conditions and VoC, both of which increase transmission, as mentioned by Campbell et al. (68). We tested this assumption by removing weather conditions from our model and found an increased transmissibility of 43%, see Appendix G for VoC. Finally, we kept the weather conditions in our model because they greatly improved the regression fit.

We emphasize that our strategy does not allow us to estimate the direct effects of NPIs, weather, and new VoC on transmission rates, but is only suggestive of associations. Indeed, the only data available are observational, and the introduction of NPI is certainly not random, but clearly depends on the state of the epidemic. To estimate the direct effect, we would need to use either methods of causal inference with time-varying confounders or dynamic causality theory (72). In the latter case, it is possible to use mechanistic models to estimate direct effects even in observational studies. However, transmission rates would have to be estimated directly in a one-step procedure using a parametric function that depends on these effects. This often relies on strong assumptions, notably the parametric shape of the transmission rate over time. But determining the shape of this dynamic is precisely the interest of this work using Kalman filters.

Overall, this work is one of the first attempts to retrospectively assess the associations between multiple NPIs and transmission rates over a one-year period of the COVID-19 epidemic. In addition to applying a novel methodology to a current and important application, this work could be extended to generate “what-if” scenarios and help determine appropriate NPI implementations for future waves of infection.

The authors thank the `opencovid-19` initiative for their contribution to the opening of the data used in this article. This work is supported in part by the Inria Mission COVID19, project GESTEPID. The authors sincerely thank Jane Heffernan for scientific discussions and thorough proofreading of the article. We also thank Linda Wittkop, Jane Heffernan, Quentin Clairon, Thomas Ferté, and Maria Pietro for constructive discussions about this work. Part of the experiments presented in this paper were carried out using the PlaFRIM experimental testbed, supported by Inria, CNRS (LABRI and IMB), Université de Bordeaux, Bordeaux INP and Conseil Régional d’Aquitaine (see https://www.plafrim.fr).

## Data Availability

All data used are available from the data.gouv.fr website of the French government.

https://www.data.gouv.fr/fr/

## Author contributions

AC, BPH, PM, MP and RT designed the study. LL and CV analyzed the data. AC, PM and MP implemented the software code. AC, BPH and MP interpreted the results. AC, BPH, LL, PM and MP wrote the manuscript.

## A Supplementary figures for hospitalization, NPIs, variants of concern and vaccination data

Figure 8 represents the elsewhere published (SI-VIC database) and publicly available data on prevalence and incidence of hospitalization for COVID-19 in 12 non-insular French regions. Representation of NPIs in all regions is available in Figure 9. Finally, representations of the VoC proportion and the vaccination coverage ramping up (1st dose) in the population in each region over time are given in Figure 10.

**Figure 8.**
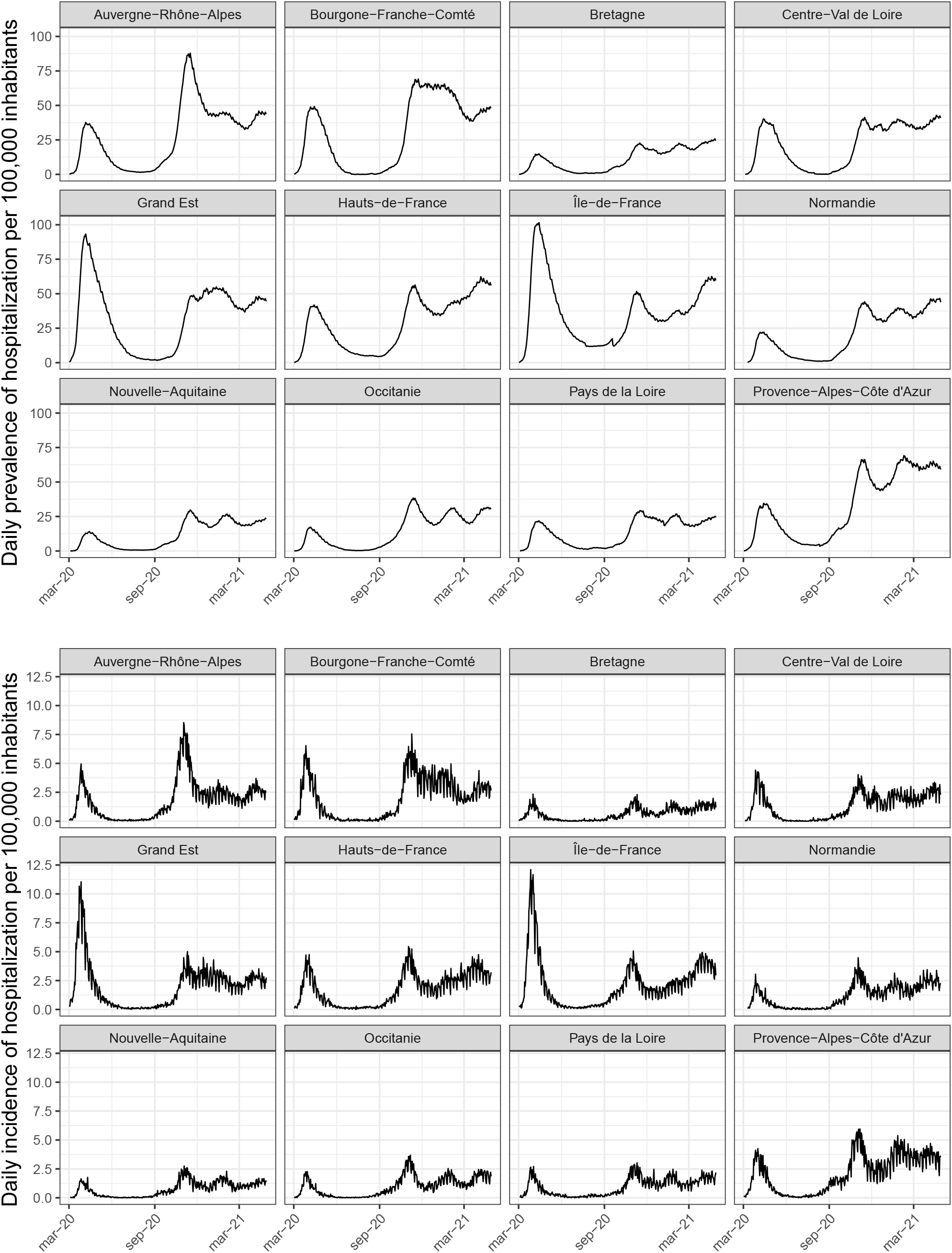
Top: total number of hospital bed occupied per 100,000 inhabitants (100, 000*/N × Y* ^*H*^). Bottom: daily number of new hospitalizations per 100,000 inhabitants 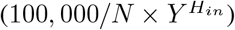.

**Figure 9.**
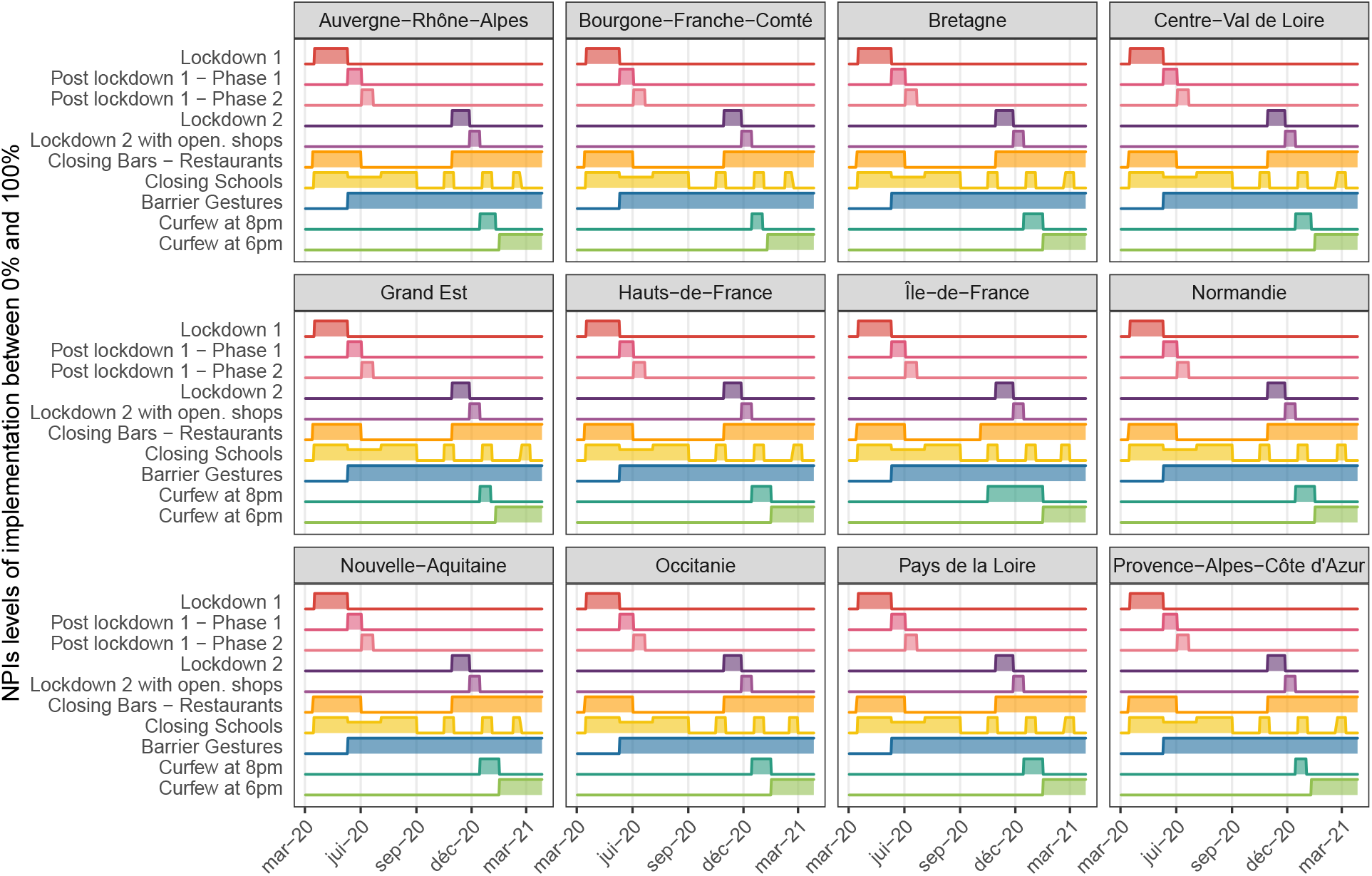
Implementation of NPIs in all regions. Differences are only impacting school closure and curfew by few days.

**Figure 10.**
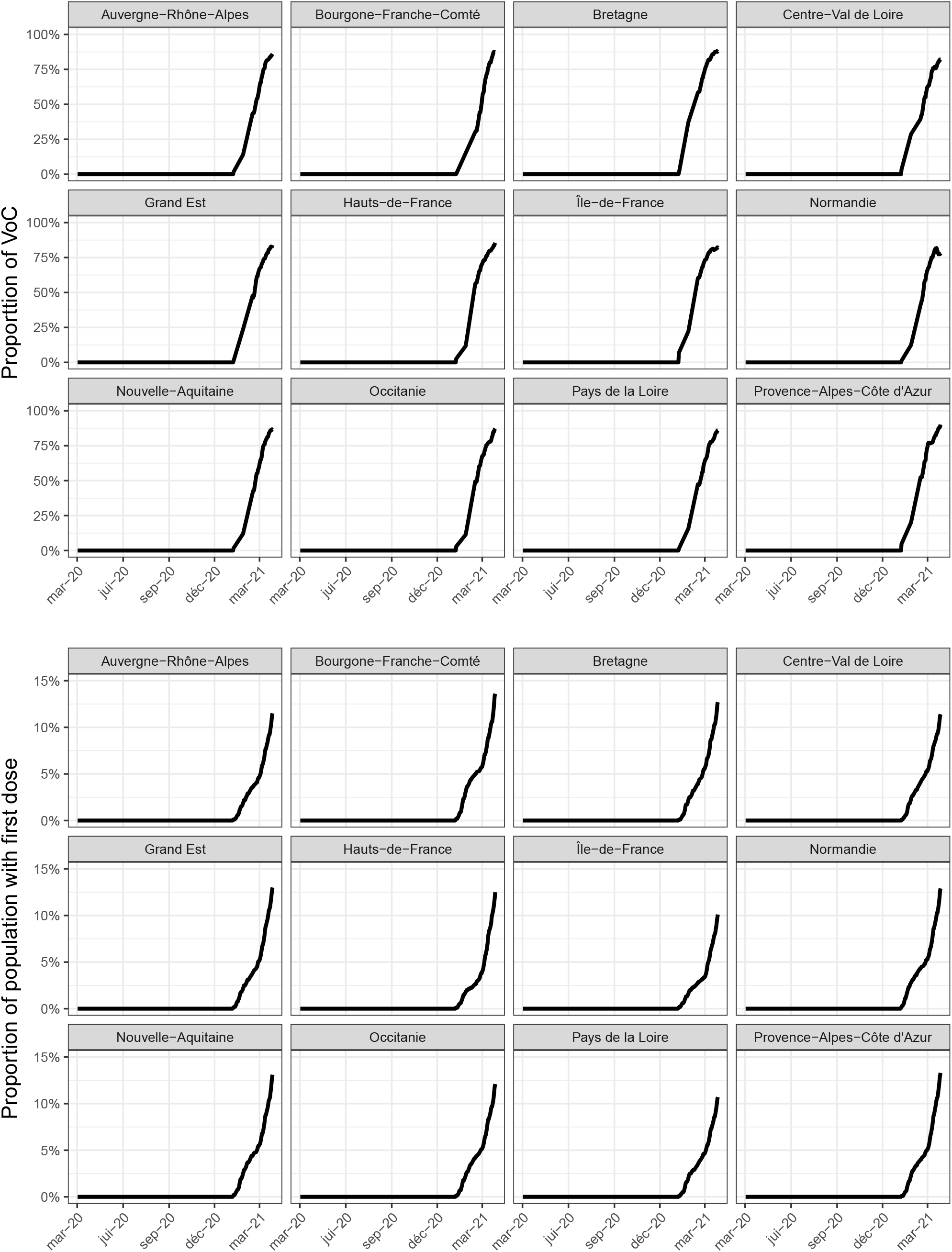
Top: Percentage of either UK, SA or BR VoC over time. Bottom: Percentage of people who have received a first dose of vaccine.

## B Estimation of the hospitalization period using the correlation between the total number of individual hospitalized daily and the daily incident number of hospitalization

The relation between the total number of individual hospitalized daily denoted by *H* and the daily incident number of hospitalization *H*_*in*_ is governed by

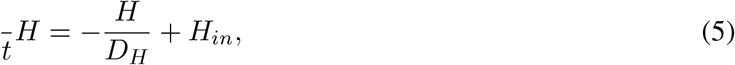

with *D*_*H*_ the hospitalization period.

Using the data of daily incident number of hospitalization 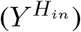 and the total number of individuals hospitalized daily (*Y* ^*H*^) over a period of about a year (391 days from March 2, 2020 to March 28, 2021), we estimate *D*_*H*_ in each region by using a mean squared estimation. The obtained values are given in Table 3. In the SEIRAH model, we fix *D*_*H*_ at the mean value.

**Table 3.** Estimation of *D*_*H*_ for the 12 regions: Île-de-France (IDF) ; Centre-Val de Loire (CVL) ; Bourgone-Franche-Comté (BFC) ; Normandie (Nor) ; Hauts-de-France (HDF) ; Grand Est (GE) ; Pays de la Loire (PL) ; Bretagne (Bret) ; Nouvelle-Aquitaine (NA) ; Occitanie (Occ) ; Auvergne-Rhône-Alpes (AURA) ; Provence-Alpes-Côte d’Azur (PACA).

|  | IDF | CVL | BFC | Norm. | HDF | GE | PL | Bret. | NA | Occ. | AURA | PACA | Nat. avg. |
| --- | --- | --- | --- | --- | --- | --- | --- | --- | --- | --- | --- | --- | --- |
| $D_H$ (days) | 18.3 | 19.5 | 18.0 | 20.6 | 18.6 | 17.7 | 16.7 | 19.6 | 18.1 | 17.4 | 17.0 | 18.1 | 18.3 |

## C Computation of the effective reproductive ratio

To compute the reproductive ratio *R*_eff_ of our SEIRAH model

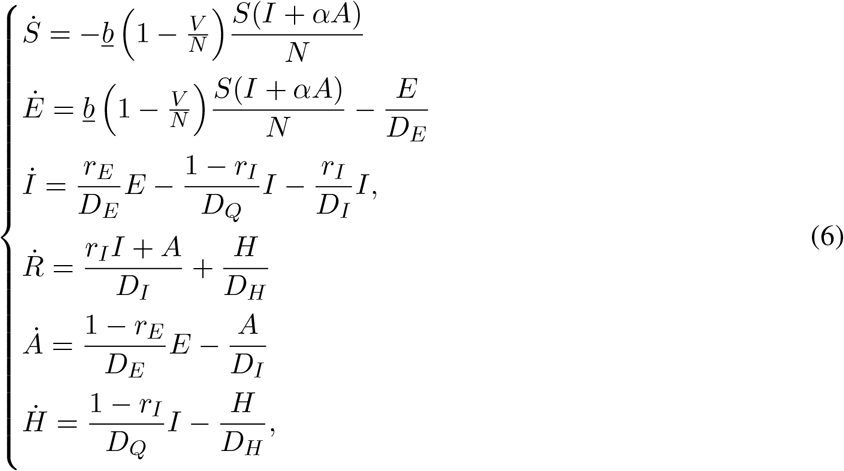

we apply the *Next Generation Matrix* approach (73). The principle consists in focusing on three categories: i) latent *E*, ii) ascertained infectious *I* and iii) unascertained infectious *A* with the following dynamics

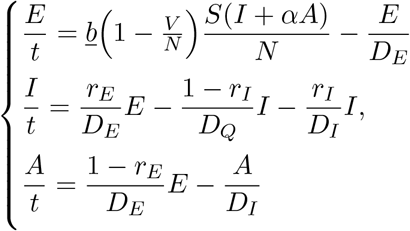

Then, we build two matrices corresponding to: (1) *V* following the arrivals and departures from one other category and (2) *F* following the arrivals from another compartment exterior to the three categories. We have

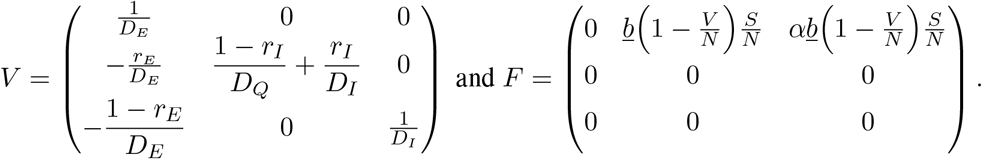

It is then well known – see for instance Perasso et al. (74) for a proof – that

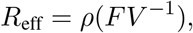

where *ρ*(*FV* ^−1^) is the spectral radius of the Next Generation Matrix *FV* ^−1^. Here, we have

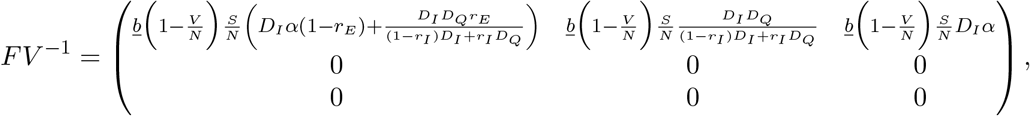

with

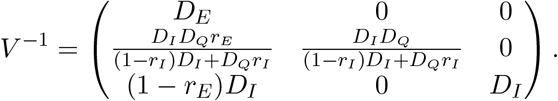

We therefore obtain

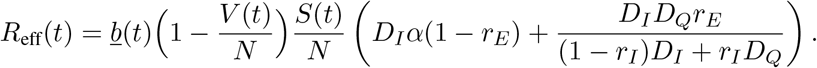

## D Initial transmission rate and attack rate estimated using our population-based Kalman filter

Table 4 shows the estimation of the initial values for the transmission rate at the regional level.

**Table 4.** Estimation of *b*_*init*_ for the 12 regions: Île-de-France (IDF) ; Centre-Val de Loire (CVL) ; Bourgone-Franche-Comté (BFC) ; Normandie (Nor) ; Hauts-de-France (HDF) ; Grand Est (GE) ; Pays de la Loire (PL) ; Bretagne (Bret) ; Nouvelle-Aquitaine (NA) ; Occitanie (Occ) ; Auvergne-Rhône-Alpes (AURA) ; Provence-Alpes-Côte d’Azur (PACA).

|  | IDF | CVL | BFC | Norm. | HDF | GE | PL | Bret. | NA | Occ. | AURA | PACA | Nat. avg. |
| --- | --- | --- | --- | --- | --- | --- | --- | --- | --- | --- | --- | --- | --- |
| $b_{init}$ | 0.789 | 0.767 | 0.784 | 0.773 | 0.781 | 0.809 | 0.761 | 0.765 | 0.768 | 0.789 | 0.786 | 0.778 | 0.779 |

## E Comparison of obtained attack rates with other studies

Our attack rates are compared to i) those obtained by Hoze et al. (62) (see Table 5), and to ii) 3 seroprevalence studies (63; 64; 65) (see Table 6).

**Table 5.** Comparison of the estimated attack rates obtained in Hoze et al. (62) (first line) with our estimated attack rates (second line) at 3 dates for the metropolitan France.

|  | May 11, 2020 | October 31, 2020 | January 15, 2021 |
| --- | --- | --- | --- |
| Hoze et al.(62) | 5.7% [5.1% ; 6.4%] | 11% [9.7% ; 12.4%] | 14.9% [13.2% ; 16.9%] |
| Proposed estimates | 5.69% [5.61% ; 5.77%] | 12.78% [11.98% ; 13.66%] | 18.92% [16.76% ; 21.43%] |

**Table 6.** Comparison of the estimated attack rates obtained in 3 seroprevalence studies. * (63; 64; 65) (first line). ** with our estimated attack rates (second line) averaged during the 3 corresponding date intervals.

|  | May 2 - June 2, 2020 | May 4 - June 23, 2020 | Oct. 5 - Oct. 11, 2020 |
| --- | --- | --- | --- |
| Auvergne-Rhône-Alpes* | 4.8% | - | - |
| Auvergne-Rhône-Alpes** | 4.48% | 4.54% | 7.15% |
| Bourgogne-Franche-Comté* | 1.5% | - | 9.3% |
| Bourgogne-Franche-Comté** | 6.04% | 6.14% | 9.33% |
| Bretagne* | 3.1% | - | - |
| Bretagne** | 1.75% | 1.79% | 3.51% |
| Centre-Val de Loire* | 2.1% | - | - |
| Centre-Val de Loire** | 5.13% | 5.26% | 8.15% |
| Grand Est* | 6.7% | 9% | 11.6% |
| Grand Est** | 10.85% | 11.36% | 12.72% |
| Hauts-de-France* | 2.9% | - | - |
| Hauts-de-France** | 5.42% | 5.64% | 8.64% |
| Île-de-France* | 9.2% | 10% | 14.8% |
| Île-de-France** | 12.57% | 13.06% | 17.97% |
| Nouvelle-Aquitaine* | 2.0% | 3.1% | - |
| Nouvelle-Aquitaine** | 1.51% | 1.52% | 3.06% |
| Normandie* | 1.9% | - | - |
| Normandie** | 2.28% | 2.31% | 5.25% |
| Occitanie* | 1.9% | - | - |
| Occitanie** | 1.71% | 1.74% | 5.17% |
| Provence-Alpes-Côte d’Azur* | 5.2% | - | - |
| Provence-Alpes-Côte d’Azur** | 4.49% | 4.63% | 8.55% |
| Pays de la Loire* | 3.4% | - | - |
| Pays de la Loire** | 2.62% | 2.69% | 4.01% |

## F Regression model

The model writes as follow for each region *i* ∈ 1 … 12:

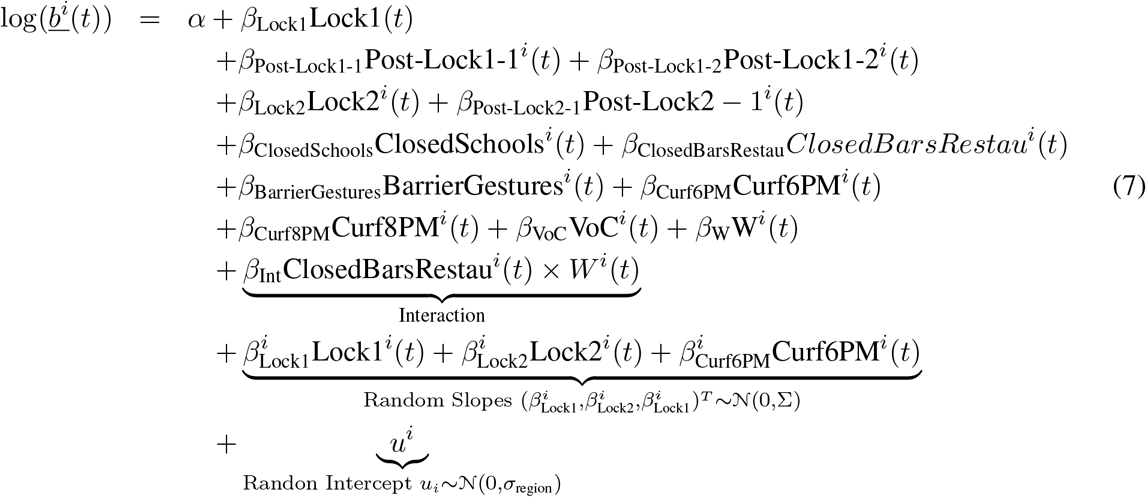

Regression residuals, fixed and random effects are for the selected model given in Eq. (8) are given in Table 7. Covariance matrix is given in Table 8.

**Table 7.** Scaled residuals, random and fixed effects of regression model (8)

| Scaled residuals |  |  |  |  |  |
| --- | --- | --- | --- | --- | --- |
|  | Min | 1Q | Median | 3Q | Max |
|  | -3.9394 | -0.4817 | -0.0033 | 0.5609 | 10.5139 |
| Random effects |  |  |  |  |  |
|  | Variance | Std.Dev. | Corr |  |  |
| $\sigma_{\text{region}}$ | 0.005159 | 0.07183 | | | |
| $\sigma_{\text{Lock1}}$ | 0.106598 | 0.32649 | -0.43 | | |
| $\sigma_{\text{Lock2}}$ | 0.018070 | 0.13443 | 0.35 | -0.52 | |
| $\sigma_{\text{Curf-6PM}}$ | 0.002827 | 0.05317 | -0.29 | 0.12 | -0.39 |
| Fixed effects |  |  |  |  |  |
|  | Estimate | Std. Error | df | t value | Pr(> \|t\|) |
| $\alpha$ | -0.40919 | 0.02595 | 25.34463 | -15.766 | 1.30e-14 |
| Lock1 | -1.52327 | 0.09631 | 11.76340 | -15.817 | 2.75e-09 |
| Post-Lock1 - 1 | -0.77153 | 0.02037 | 4620.46129 | -37.869 | < 2e-16 |
| Post-Lock1 - 2 | -0.65810 | 0.01427 | 4634.29686 | -46.120 | < 2e-16 |
| Lock2 | -0.76626 | 0.04312 | 13.69861 | -17.771 | 7.47e-11 |
| Lock2 - red. | -0.71299 | 0.02169 | 4644.40638 | -32.876 | < 2e-16 |
| Closed Schools | -0.07150 | 0.00866 | 4639.83981 | -8.256 | < 2e-16 |
| Closed Bars&Rest. | -0.10746 | 0.01492 | 4623.96873 | -7.201 | 6.93e-13 |
| Barrier Gestures | -0.61300 | 0.01963 | 4652.90602 | -31.229 | < 2e-16 |
| Curf. 6PM | -0.35386 | 0.02629 | 59.55020 | -13.461 | < 2e-16 |
| Curf. 8PM | -0.32590 | 0.01951 | 4404.57885 | -16.707 | < 2e-16 |
| Variants | 0.19505 | 0.02759 | 4605.07217 | 7.071 | 1.77e-12 |
| Weather | -1.03117 | 0.04013 | 4654.84448 | -25.696 | < 2e-16 |
| Bar&Rest. : Weather | 0.11877 | 0.05706 | 4501.30235 | 2.082 | 0.0374 |

**Table 8.**
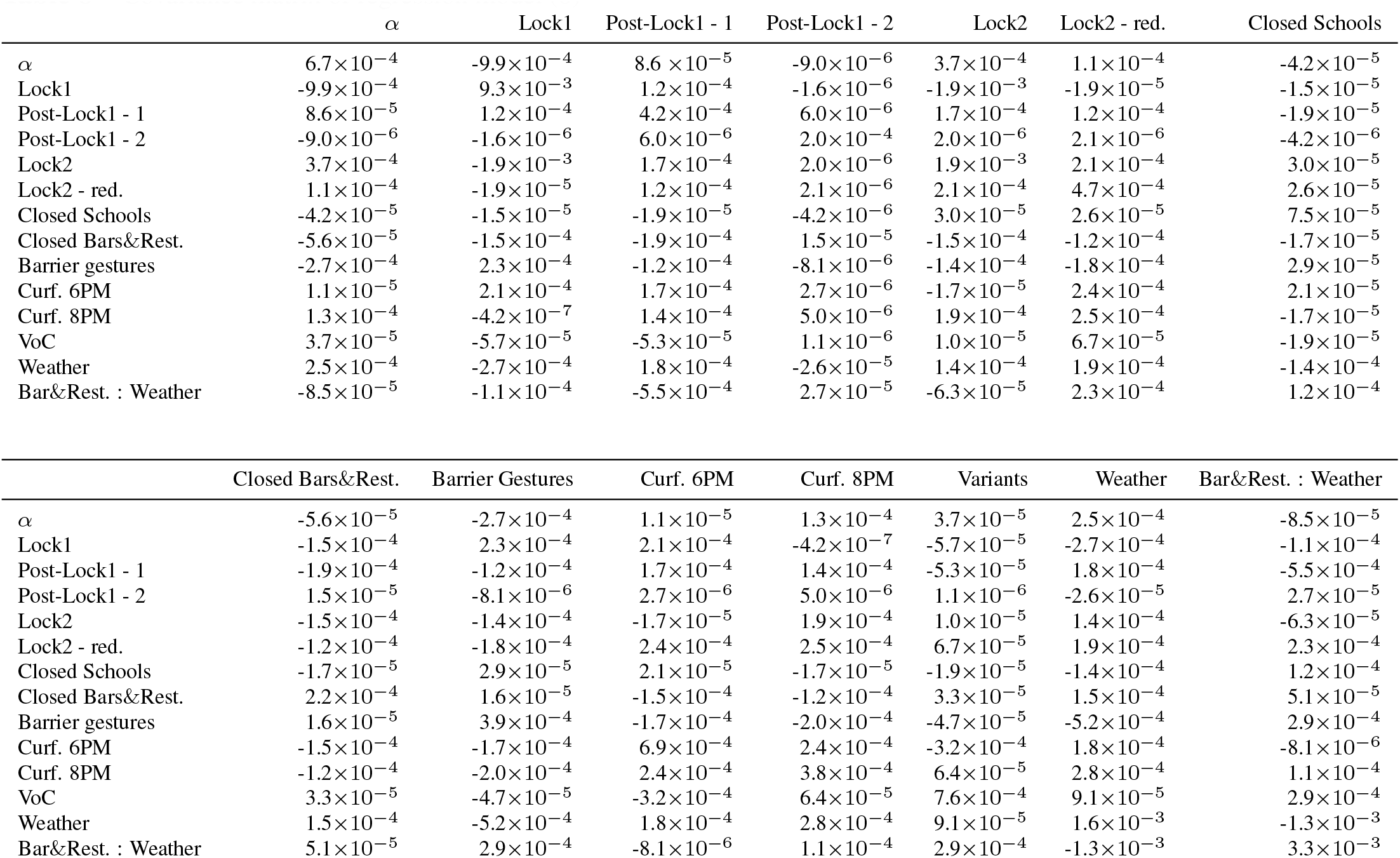
Covariance matrix of regression model (8)

## G Comparison with other regression models

In this part, we compare our regression model to other regression models. We start by considering a simple model neglecting the weather (Model 1) and then we consider a model integrating the weather variable but neglecting the interaction with the bars and restaurants (Model 2) and the selected model (Model 3). Model 4 corresponds to Model 3 but without considering the delay of 7 days after the lockdowns. Table 9 summarizes the results. Figure 11 shows the fits.

**Table 9.** Estimation of the associations between the transmission rates and the weather, the VoCs and the NPIs. Negative (resp. positive) values correspond to a decrease (resp. an increase) of the transmission rate.

| NPI/Variants/Weather | Model 1 | Model 2 | Model 3 | Model 4 |
| --- | --- | --- | --- | --- |
| Lockdown 1 (delay of 7 days) | -83% [-86% ; -80%] | -78% [-82% ; -74%] | -78% [-82% ; -74%] | -65% [-71% ; -58%] |
| Post lockdown 1 - Phase 1 | -54% [-56% ; -53%] | -53% [-54% ; -51%] | -54% [-56% ; -52%] | -45% [-49% ; -40%] |
| Post lockdown 1 - Phase 2 | -49% [-51% ; -47%] | -48% [-50% ; -47%] | -48% [-50% ; -47%] | -48% [-50% ; -46%] |
| Lockdown 2 (delay of 7 days) | -49% [-53% ; -44%] | -53% [-57% ; -49%] | -54% [-57% ; -49%] | -41% [-46% ; -35%] |
| Lockdown 2 with open. shops | -38% [-41% ; -35%] | -51% [-53% ; -49%] | -51% [-53% ; -49%] | -54% [-57% ; -51%] |
| Closing schools | -15% [-16% ; -13%] | -7% [-9% ; -6%] | -7% [-8% ; -5%] | -3% [-6% ; -1%] |
| Closing bars & restaurants | 4% [1% ; 7%] | -10% [-13% ; -8%] | -10% [-13% ; -8%] | -24% [-29% ; -19%] |
| Barrier gestures | -63% [-64% ; -61%] | -46% [-48% ; -44%] | -46% [-48% ; -44%] | -36% [-40% ; -31%] |
| Curfew at 6PM | -18% [-22% ; -13%] | -30% [-33% ; -26%] | -30% [-33% ; -26%] | -28% [-34% ; -23%] |
| Curfew at 8PM | -5% [-8% ; -2%] | -28% [-31% ; -25%] | -28% [-31% ; -25%] | -33% [-37% ; -28%] |
| Proportion of variants | 44% [36% ; 52%] | 20% [14% ; 27%] | 22% [15% ; 28%] | 6% [-2% ; 15%] |
| Seasonal weather conditions | - | -63% [-65% ; -60%] | -64% [-67% ; -61%] | -73% [-76% ; -69%] |
| Weather cond./closing bars & rest. | - | - | 13% [1% ; 26%] | -32% [-42% ; -19%] |
| AIC | -707 | -1486 | -1485 | 2224 |

**Figure 11.**
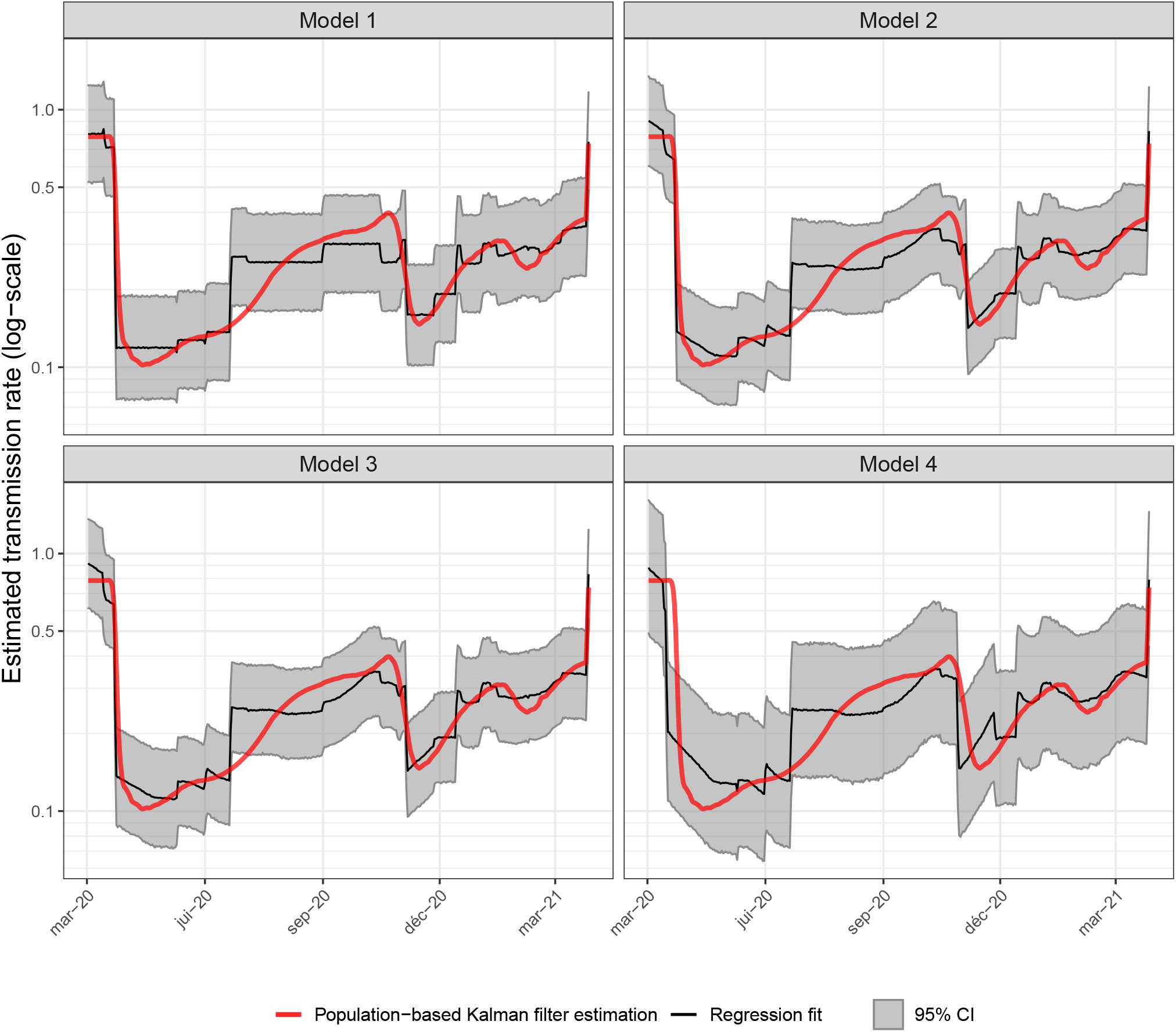
Results of regression models of the transmission rate obtained with the population based Kalman filter using Model 3 given in Equation (8). Top-Left: Mean fit with linear model without considering weather effect (Model 1). Top-Right: Mean fit with linear model considering linear weather effect (Model 2). Bottom-Left: Mean fit with linear model with linear weather effect and with closed bar and restaurants and weather interaction (Model 3). Bottom-Right: Mean fit with linear model with linear weather effect and with closed bar and restaurants and weather interaction (Model 4) but without taking into account the delay of 7 days after the lockdowns. Random effects are considered on the intercept, the lockdowns and the curfew at 6PM.

Using the first model (AIC=-707), we obtain a negative association between the closure of bars and restaurants and the transmission rates which is not realistic. This is due to the fact that the bars and restaurants were open during summer when the transmission rate was very low with a effective reproductive number inferior to 1 in all regions. That is why in a second model, we add the weather variable. The AIC of this model is larger superior to the first ones (AIC = -1486). The third model assumes that there is an interaction between the closures of bar and restaurants and the weather to take into account the use of terraces (which have been expanded in many places since the beginning of the pandemic). The AIC is similar to the second model (AIC = -1485). The AIC of Model 4 is very large (AIC = 2224) compared to other ones validating the delay of 7 days.

We found that the three VoCs (alpha, beta, and gamma) are ∼ 20% (Model 2 or 3) to ∼ 45% (Model 1) more transmissible than the historical lineage. The estimated value with Model 1 seems more realistic comparing to the literature and the end of the curve is better fitted. This is an important limitation of our work.

